# Saturation genome editing of *BARD1* resolves VUS and provides insight into BRCA1-BARD1 tumor suppression

**DOI:** 10.1101/2025.11.03.25339440

**Authors:** Ivan Woo, Silvia Casadei, Matthew W. Snyder, Nahum T. Smith, Sabrina Best, Malvika Tejura, Pankhuri Gupta, Abbye E. McEwen, Mason Post, Audrey Hamm, Moez Dawood, Airi Hosokai, Alicia Xu, Riddhiman K. Garge, Shawn Fayer, Terra Brannan, Marcy E. Richardson, Sriram Pendyala, Sarah Heidl, Lara Muffley, Douglas M. Fowler, Lea M. Starita

## Abstract

*BARD1* encodes a dimeric partner of BRCA1 and is required for homology-directed double strand DNA break repair^1–5^. Germline loss-of-function variants in *BARD1* are linked to elevated risk for breast cancer and neuroblastoma^6–8^ and PARP inhibitors have shown effectiveness against BARD1-deficient tumors. However, the majority of *BARD1* variants identified by genetic testing for cancer risk are variants of uncertain significance (VUS)^9^ limiting clinical utility. We used saturation genome editing^10^ to assess the impact of nearly 11,000 single-nucleotide variants and three base-pair deletions across all 11 coding exons of *BARD1* on cellular fitness and gene expression. The cellular fitness data are nearly perfectly concordant with known pathogenic and benign *BARD1* variants^9^ (AUC > 0.99) and loss-of-function missense variants in all three functional domains were associated with elevated risk for breast cancer^6,8^. When used for clinical variant classification, our data resolved 95.4% of existing *BARD1* variants of uncertain significance. Comparison of cellular fitness data to known structures^11–14^, further solidifies that BARD1’s role in homology-directed repair is required for tumor suppression in humans^15^. These results will immediately improve clinical genetic testing and decision-making for patients with *BARD1* variants and deepen our understanding of BARD1’s role in maintaining genome integrity.

## Main

The BRCA1-Associated RING Domain protein, BARD1 [OMIM 601593], is required for the BRCA1 tumor suppressor to fold and function^1–5^. *BARD1* variants have been linked to hereditary breast cancer and neuroblastoma through case-control studies^6,7^. Accordingly, *BARD1* is included on genetic testing panels for cancer risk. ClinVar^9^ contains 3,725 unique single nucleotide variants (SNVs) and 29 three base-pair (bp) deletions of *BARD1*, with 61.4% (2,289) of SNVs and 65.5% (19) 3-bp deletions classified as variants of uncertain significance (VUS) or have conflicting interpretations. Among missense variants, 97.7% (2,077 of 2,126) are VUS or have conflicting interpretations. Because germline pathogenic *BARD1* variants confer moderate 17–30% lifetime breast cancer risk^6^, enhanced screening is recommended for heterozygous individuals^16^. Additionally, poly (ADP-ribose) polymerase (PARP) inhibitors are FDA approved for BARD1-deficient breast cancers^17^ and have shown efficacy for neuroblastoma^18^. Thus, our inability to understand how missense variation affects BARD1 function, and classify variants accordingly, limits cancer risk assessment and treatment guidance for many individuals.

BARD1, like BRCA1, is required for repairing double-strand DNA breaks by homology-directed repair (HDR)^3,19,20^ and mediates other diverse molecular functions through multiple multiple partners^1,21–24^. BARD1 protein interactions are largely mediated through three folded domains: an N-terminal RING domain, a central ankyrin repeat domain (ARD), and C-terminal tandem BRCT repeat domain. The BRCA1-BARD1 RING domains’ E3 ubiquitin ligase activity promotes HDR through histone H2A ubiquitylation, preventing 53BP1 binding at double-strand breaks and thus promoting HDR over non-homologous end joining (NHEJ)^19^. The ARD recognizes unmethylated histone H4K20me0 tails marking newly replicated chromosomes^25^, thereby ensuring that HDR occurs only during the G2 phase of the cell cycle. The BRCT domain recognizes DNA damage-associated histone H2AK13ub/K15ub marks^12,14,19^, recruiting the BRCA1-BARD1 dimer to DNA damage sites. While these and other BARD1 interaction partners have been identified, we lack granular understanding of which residues and specific interactions BARD1 requires for HDR.

Comprehensive variant effect maps could resolve the need for functional data to interpret BARD1 VUS and refine understanding of BARD1’s protein interactions and functions. Multiplexed assays of variant effect effect (MAVEs) can furnish such maps by measuring the functional impact of all possible single nucleotide or amino acid substitutions^26,27^, revealing key domains and residues required for protein function, delineating roles in protein stabilization versus interactions, and uncovering disease mechanisms.

Additionally, MAVE-derived functional data can drive the reclassification of more than half of VUS^28^. Saturation genome editing (SGE) is a MAVE approach where sequence variants are edited into the endogenous genome of a cell line, allowing assessment of functional impact on cell survival with nucleotide resolution^10,29,30^. This captures effects on protein function, RNA expression, and splicing. SGE of essential genes in haploid cells has been used to assess variants in *BRCA1*^10,31^, *DDX3X*^32^, *VHL*^33^, *BAP1*^34^, *BRCA2*^35^, and *RAD51C*^36^—discriminating pathogenic from benign variants with high fidelity. *BARD1*, like *BRCA1* and other HDR pathway members, is essential in HAP1 cells and amenable to SGE^37^ (**Fig. 1a**).

**Figure 1.**
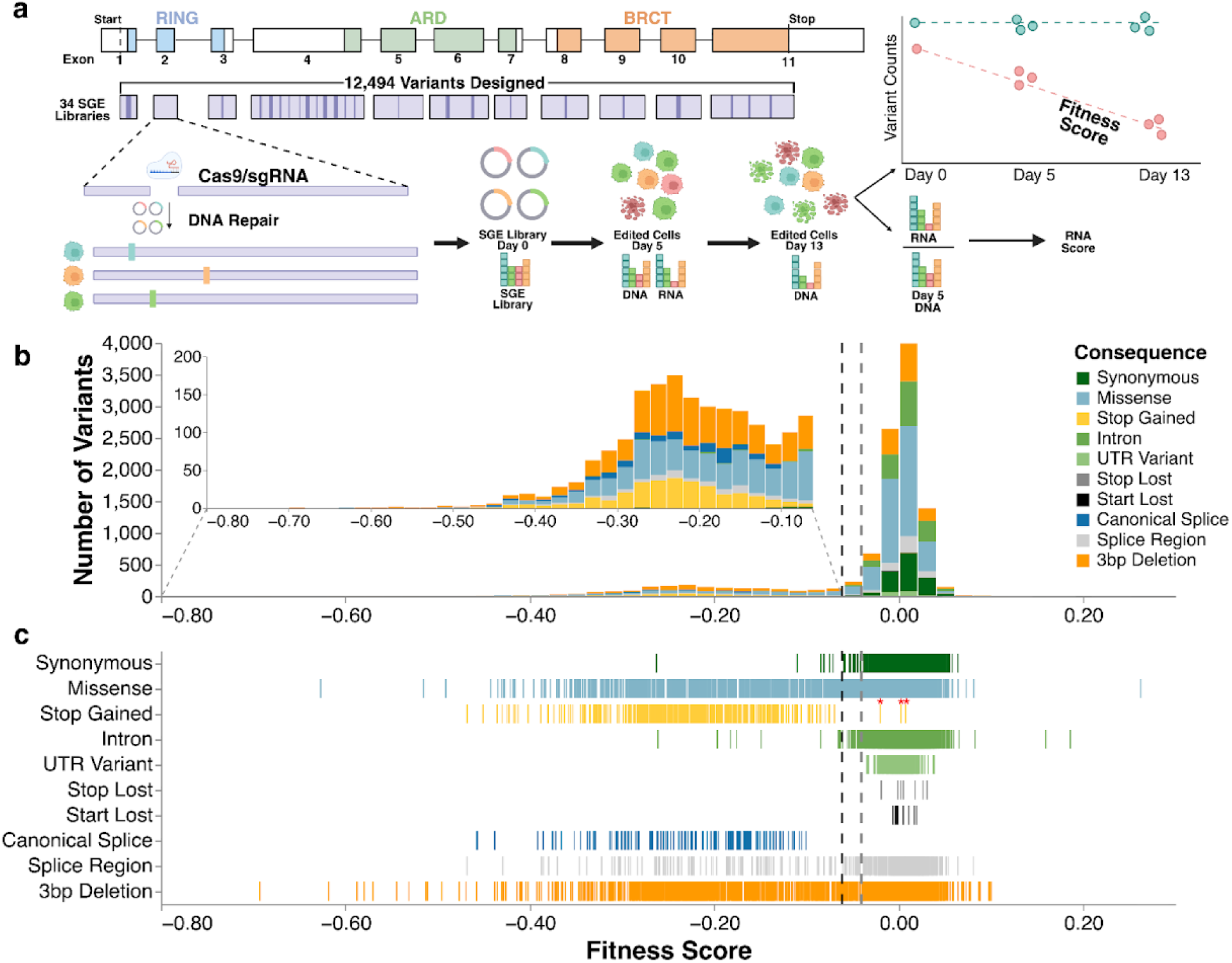
Saturation Genome Editing (SGE) of *BARD1* generates variant effect scores for 10,915 variants. (a) A map of the exon structure of *BARD1*, followed by the coverage of the 34 repair template libraries required to saturate all coding exons and proximal introns. Many exons that are too long for a single repair template library require sub-exon level repair template libraries and guide RNAs. For example, full coverage of exon 4 required sub-libraries A-L and 12 different guides. A schematic shows the SGE experimental strategy and scoring process. (b) A histogram of fitness scores (n = 10,915). Variants are colored by mutational consequence as annotated by VEP^67^. The black dotted vertical line indicates the threshold for the LoF class at −0.0615. Gray dotted vertical line indicates the threshold for the functionally normal class at −0.0406. The inset highlights LoF variants. (c) Strip plot of fitness scores organized by mutational consequence (n = 10,915). The black and gray vertical dotted lines represent thresholds for the LoF and functionally normal classes.

Here, we measured the effect of 8,818 *BARD1* SNVs and 2,097 3-bp deletions on cell survival and 6,384 SNVs on mRNA expression using SGE. Loss-of-function (LoF) missense variants and deletions were enriched in folded domains relative to disordered domains. Our SGE data discriminated between pathogenic and benign variants with near-perfect accuracy (AUC = 0.99), correctly identifying the only confirmed pathogenic missense variant, p.Cys71Tyr, as LoF. LoF missense variants in the ARD and BRCT domains were enriched in breast cancer cohorts, linking these domains’ molecular functions to BARD1 tumor suppression. Our data provided strong evidence for variant classification, enabling resolution of 95.4% of VUS when functional evidence was applied. The nucleotide resolution of SGE enabled us to discern splice and RNA expression effects from protein effects, revealing that the annotated start codon is dispensable and that several synonymous and intronic variants are LoF. Analysis of missense variation highlighted key residues for known protein interactions and identified a previously unknown interaction surface. While BARD1 has been ascribed multiple cellular roles, we provide additional evidence that its HDR function is linked to cancer risk. This comprehensive variant effect dataset immediately informs inherited cancer risk assessment and treatment decisions.

### Fitness scores for 8,818 SNVs and 2,097 3-base-pair deletions for *BARD1*

To perform SGE on all 2,334 bases of *BARD1*’s coding sequence and at least 10 bases of intronic or UTR sequence surrounding coding exons, we generated 34 sgRNAs and repair template libraries encoding all possible 9,351 SNVs and approximately 2,180 unique 3-bp deletions. For each of the 34 SGE regions, we transfected a sgRNA and matching repair template library into HAP1-LIG4Δ-Cas9 cells^32^ to induce editing by HDR. All editing experiments were performed in triplicate, with editing rates that generated usable sequences (*e.g.* having all required edits) ranging from 10.5% to 39.8% across SGE regions (**Extended Data Fig. 1a**). We PCR amplified and sequenced the edited SGE regions from genomic DNA at days 5 and 13 to identify and count variants to calculate fitness scores, and from mRNA at day 5 to calculate RNA scores. Variant counts were generally reproducible across individual replicates (median Pearson’s r = 0.87) (**Extended Data Fig. 1b-c**).

We calculated fitness scores (log_2_ fold change in variant abundance per day) for 95% of all possible SNVs (8,818) and 3-bp deletions (2,097) using a continuous-time linear regression model across time points and replicates (**Fig. 1a, Extended Data Fig. 2, and Supplementary File 1**). Synonymous and intronic variants, which mostly do not impact function, had scores near zero (**Fig. 1b-c**). Nonsense and canonical splice variants, which are expected to disrupt expression and protein function, were depleted from the cell population and had negative scores, confirming that BARD1 function is required for HAP1 cell survival. Most fitness scores were near zero with a long tail of negatively scoring variants that depleted over the course of the experiment. We used the distribution of synonymous/intronic variants and nonsense variants to determine thresholds to classify variants as functionally normal or LoF using a Gaussian Mixture Model (**Fig. 1c and Extended Data Fig. 3**). As expected, 365 of 368 (99.2%) nonsense variants were LoF and 1,286 of 1,306 (98.5%) synonymous variants were functionally normal. 94.4% of the 180 canonical splice variants were LoF. In sequence elements where variants have varying effects, such as splice regions, scores were more widely distributed with 84.8% classified as functionally normal and 12.8% as LoF (**Fig 1c**). Missense variants showed a similar pattern (84.6% functionally normal, 12.7% LoF), suggesting that HAP1 cells are sensitive to loss of specific BARD1-dependent mechanisms required for cell survival.

### SGE reveals residues required for BARD1 function in the RING, ARD and BRCT domains

BARD1 has three folded domains: an N-terminal RING domain, an ankyrin repeat domain (ARD), and a C-terminal tandem BRCT repeat domain. A disordered domain encoded by exon 4 connects the RING and ARD, while a disordered domain encoded by exons 7-8 connects the ARD and BRCT (**Fig. 2a**)^14,38^. We validated that SGE scores reflect known functional effects by comparing them to results of orthogonal biochemical assays for HDR activity^39,40^, sensitivity PARP inhibitor sensitivity^19^, and *in vitro* ubiquitylation^12,41^ (**Extended Data Fig. 5a**). Fitness scores were highly concordant with these assays, demonstrating that our cellular fitness readout integrates known BARD1-dependent molecular and cellular functions.

**Figure 2.**
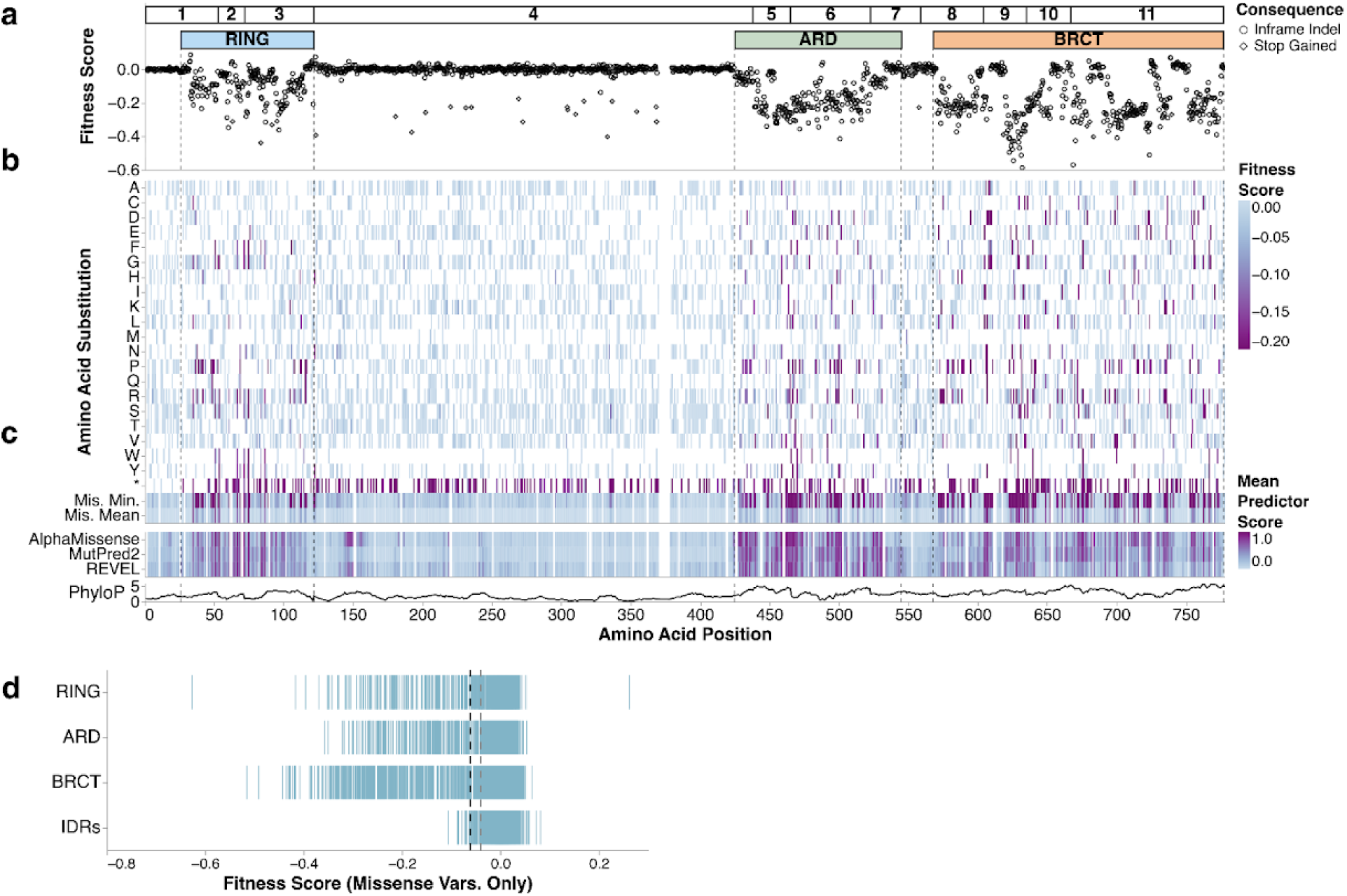
SGE identifies critical domains and functional residues of BARD1. (a) Map of programmed 3-bp deletions (n = 2,097). Amino acid positions in BARD1 are on the X-axis and fitness scores are on the Y-axis. In-frame deletions and deletions that cause stop-gain variants are denoted by shape. (b) A heatmap of fitness scores for missense and stop-gained variants. Amino acid positions in BARD1 are on the X-axis and amino acid and stop-gained (*) changes are indicated on the Y-axis. Fitness scores are colored as indicated. (c) Heatmap of minimum and mean fitness scores across missense variants and mean variant effect predictor scores. For predictors, high scores denote predicted pathogenicity, while low fitness scores indicate LoF. The bottom-most track is a line plot depicting the rolling mean phyloP score across a 10 amino acid residue window. (d) A strip plot of fitness scores for missense variants grouped by protein domain. Black and gray dashed vertical lines indicate functional class thresholds for functionally abnormal and functionally normal respectively. IDRs denote the exon 4-encoded disordered domain.

Within the exon 4 disordered domain, only 2 of 575 3-bp deletions depleted (excluding stop-gained), and large incidental in-frame deletions persisted throughout the experiment, whereas such deletions depleted in folded domain-encoding exons (**Extended Data Fig. 4**). Of 1,802 missense SNVs in the disordered domain, only 11 (0.61%) scored as LoF (**Fig. 2b, d**). Altogether, this suggests that although the exon 4-encoded region binds DNA and promotes end resection^14,38^, few single amino acids are necessary for critical BARD1 function. Notably, one exon 4-encoded LoF variant, p.Lys140Asn, has been shown to play a role in BRCA1-BARD1-catalyzed end resection and RAD51 loading^42^.

In contrast, 98.1% (576 of 587) of LoF missense variants overlapped the three folded domains, representing a 36-fold enrichment relative to disordered domains (p = 2.8e-128, Fisher’s exact test) (**Fig. 2b, d**). The folded domains showed similar missense sensitivity, with 20.1%, 21.9%, and 23.5% of variants in the RING, ARD, and BRCT showing LoF respectively. Pairwise comparisons revealed no significant differences between domains (Fisher’s exact tests, adjusted p > 0.05), and experimental replicates showed high correlation (median Pearson’s r = 0.89) (**Extended Data Fig. 2**).

Comparison to computational predictors AlphaMissense^43^, MutPred-2^44^, and REVEL^45^ revealed similar patterns but a tendency to overpredict pathogenicity (**Fig. 2c**, **Extended Data Fig. 5b**). Incorrectly predicted variants with at least moderate pathogenicity evidence^46^ were enriched in the ARD for all three predictors. Additionally, AlphaMissense predicted variants at residues 140–160 to be pathogenic and CADD^47^ predicted start-loss variants to be deleterious, but both had normal fitness scores (**Fig. 2c**, **Extended Data Fig. 5b**).

### SGE reveals germline BARD1 variants that are linked to increased cancer risk

To assess relevance to human disease, we compared our SGE data, generated in a haploid human cell line, to human genetics databases. LoF BARD1 variants had significantly lower minor allele frequency (MAF) than functionally normal variants in gnomAD^48^ (median MAF 6.2e-7 vs. 1.2e-6, p = 2.3e-7) and the Regeneron Million Exome Variant Browser^49^ (6.1e-7 vs. 1.2e-6, p = 0.001). This suggests that variant effects measured by SGE are relevant to human health as variants not tolerated in SGE are also infrequent in the human population (**Extended Data Fig. 6**).

Fitness scores of pathogenic or likely pathogenic (PLP) and benign or likely benign (BLB) *BARD1* variants in ClinVar had different distributions (mean scores −0.23 vs. 0, p = 6.4e-24) (**Fig. 3a**). Of variants scored by SGE, 202 are classified as PLP in ClinVar (139 stop-gained, 58 canonical splice, 1 missense, 4 other). SGE correctly identified 99% (199 of 202) as LoF, including the only bonafide pathogenic missense variant p.Cys71Tyr. Moreover, we correctly classified 99% (987 of 1,001) of BLB variants (711 synonymous, 134 splice-region, 40 missense, 116 other) as functionally normal, with eleven variants falling in the indeterminate range. This high sensitivity and specificity (AUC = 0.99, **Fig. 3b**) demonstrates that SGE accurately predicts disease-relevant variant effects.

**Figure 3.**
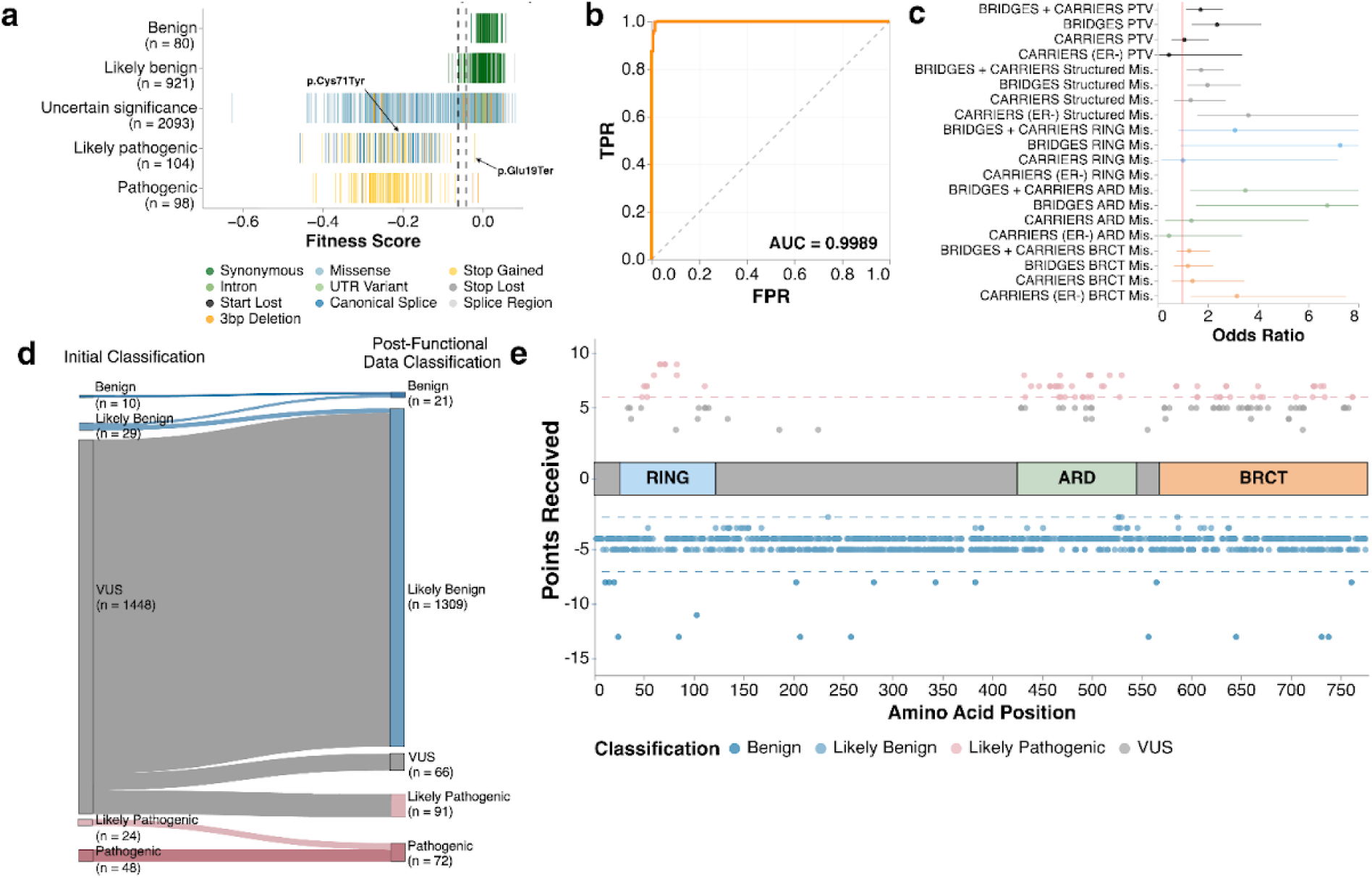
Comparison of *BARD1* fitness scores to human genetics studies. (a) A strip of *BARD1* fitness scores by ClinVar label (n = 3,296). All single nucleotide variants and 3-bp deletions accessed from ClinVar^9^ on September 12, 2025 and had at least 1-star review status. (b) Receiver operator characteristic (ROC) curve for the ability of BARD1 fitness scores to discriminate pathogenic/likely pathogenic from benign/likely benign variants from ClinVar. (c) Odds ratios for the occurrence of LoF variants in breast cancer cases vs. controls in the listed cohorts for either all protein or specified domain. For all cohorts, only data from population-based studies was used. The vertical red line denotes an odds ratio of one. Points denote the estimated odds ratio and whiskers denote the 95% confidence interval. (d) Sankey plot of Ambry VUS before and after addition of functional evidence. (e) Scatter plot of missense variants reclassified using functional evidence. A cartoon of BARD1’s three domains is centered at Y = 0. Y-axis represents number points toward a benign (negative) or pathogenic (positive) classification. Horizontal lines denote the classifications thresholds; the pink line at Y = 6 denotes the threshold for “Likely Pathogenic”, the light blue line at Y = −2 denotes the cutoff for “Likely Benign”, and the blue line at Y = −7 denotes the cutoff for “Benign”. The X-axis represents the amino acid position of the variant. Variants are colored by their final classification.

To further validate our results, we evaluated whether LoF missense variants were enriched in the BRIDGES^6^ and CARRIERS^8^ breast cancer case-control cohorts. LoF missense variants were enriched in breast cancer cases compared to healthy controls (OR = 1.72, 95% CI = 1.19–2.48) (**Fig. 3c, Supplementary Table 1**). Notably, this odds ratio was not significantly different from those of protein-truncating variants (OR = 1.71, 95% CI = 1.13–2.59, p = 0.99, Z-test), suggesting that LoF missense and truncating BARD1 variants confer similar cancer risk. However, the odds ratios for our LoF missense variants are significantly different than the odds ratio of 1.00 (95% CI 0.89–1.12) reported in the BRIDGES study for rare missense variants in *BARD1* (p = 0.01 and p = 0.005 respectively, Z-test) suggesting that assessing the risk associated with *BARD1* missense variants has been hampered by ascertainment of LoF variants. Reinforcing the enrichment results for SGE LoF variants, missense variants having at least moderate evidence of pathogenicity by the MutPred-2 variant effect predictor were also enriched in breast cancer cases (OR = 2.64, 95% CI = 1.10–6.32) (**Extended Data Fig. 5c**)^44,46^. Moreover, utilizing estrogen receptor (ER) status annotations in the CARRIERS cohort, we found that germline LoF missense variants in structured domains were enriched in ER- breast cancer cases (OR = 3.61, 95% CI = 1.58–8.24) compared to healthy controls (**Fig. 3c**), aligning with CARRIERS^8^ findings that protein-truncating BARD1 variants were associated with elevated ER- breast cancer risk.

While the only bonafide pathogenic BARD1 missense variant, p.Cys71Tyr, is in the RING domain, we investigated whether LoF missense variants in the ARD and BRCT domains were also associated with increased disease risk. ARD LoF missense variants were enriched in the BRIDGES^6^ cohort (population-based OR = 6.75, 95% CI = 1.52–29.9), and BRCT LoF variants were associated with increased ER- breast cancer risk in the CARRIERS^8^ cohort (OR = 3.16, 95% CI = 1.33–7.51) (**Fig. 3c**). Neither cohort had sufficient variants in the smaller RING domain for statistically significant results (**Supplementary Table 1**). Additionally, missense variants that scored as functionally normal by SGE showed no statistically significant enrichment in breast cancer cases relative to healthy controls (**Supplementary Table 1**). These results demonstrate that SGE-measured variant effects are disease-relevant and that LoF missense variants across all three BARD1 functional domains are associated with increased cancer risk.

### BARD1 fitness scores enable VUS reclassification

Next, we assessed the clinical utility of these data for VUS reclassification. Calibration is required to translate fitness scores to evidence for variant classification. The current ClinGen-approved calibration method calculates OddsPath, a Bayesian odds ratio used to assign evidence based on the ability of the assay to correctly identify previously classified pathogenic and benign variants as LoF or functionally normal, respectively^50^. We collected 186 PLP and 959 BLB variants from ClinVar for calibration (**Supplementary Table 2**). We included stop-gained, splice site and synonymous variants because the fitness scores for nonsense variants overlapped LoF missense and synonymous overlapped functionally normal missense scores (**Fig. 1c**), LoF missense variants are linked to cancer risk through either family^51^ or case-control analyses, (**Fig. 3c**) and for practical purposes, there were too few classified pathogenic missense variants available to assign any benign evidence otherwise (**Extended Data Fig. 7**). The accurate positive and negative predictive values of SGE resulted in an OddsPath of 316.23 which equated to PS3_strong evidence (4 points) towards pathogenic classifications and an 1/OddsPath of 0.0054 for functionally normal, assigning strong evidence (−4 points) toward benign classifications (**Supplementary Table 2**).

To reclassify VUS we combined the additional evidence from Ambry Genetics and the calibrated functional evidence to reclassify 87.6% (1,382/1,578) of VUS in Ambry’s database according to the ACMG/AMP v3 recommendations with the exception that likely benign classifications required −2 points^52,53^. For the 1,448 VUS for which we had functional evidence, 1,382 (95.4%) were reclassified. 1,291 (89.2%) VUS were reclassified as BLB and 91 (6.3%) are reclassified as PLP and 66 (4.5%) remain VUS (**Fig. 3d, Extended Data Fig. 7, and Supplementary Table 2**). The functional evidence enabled reclassification of all missense VUS receiving benign evidence. All remaining missense VUS have points toward pathogenic classifications, of those with +4 and +5 points (‘high VUS’), all but 1 are in BARD1’s three folded domains (**Fig. 3e**).

100% of reclassifications were driven by functional evidence. Computational predictions (PP3/BP4) contributed to 96% reclassifications, population frequency (PM2_supporting) contributed to 70%, and predicted functional consequence (PVS1) contributed to 0.6%. Variants that remained VUS did not have functional evidence (n = 88), had indeterminate fitness scores (n = 42) or did not have enough evidence beyond functional evidence for reclassification (n = 66). The reclassification rate of 95.4% when functional evidence was available highlights the high clinical utility of *BARD1* SGE data.

### SGE identifies SNVs that impact BARD1 expression

To parse LoF mechanisms, we first looked for unexpected results that might reflect RNA- versus protein-level variant effects. We noticed that start-loss variants and three stop-gained variants in the first exon were tolerated. Of these variants, three start-loss variants (p.Met1Ile, p.Met1Val, and p.Met1Thr) are VUS or have conflicting classifications of pathogenicity and one stop-gained variant (p.Glu19Ter) is PLP in ClinVar. These variants occurred between the annotated start codon and p.Met26, suggesting that p.Met26 may be capable of initiating translation. Double-mutant experiments targeting both initiators confirmed that p.Met1 and p.Met26 can independently initiate BARD1 translation: when p.Met1 was lost, variants abrogating p.Met26 became deleterious (**Fig. 4a-b**). Additionally, we saw no loss of RNA expression for variants at either p.Met1 or p.Met26 (**Extended Data Fig. 8**). These results indicate that start-loss and stop-gained variants upstream of p.Met26 are unlikely to contribute to disease.

**Figure 4.**
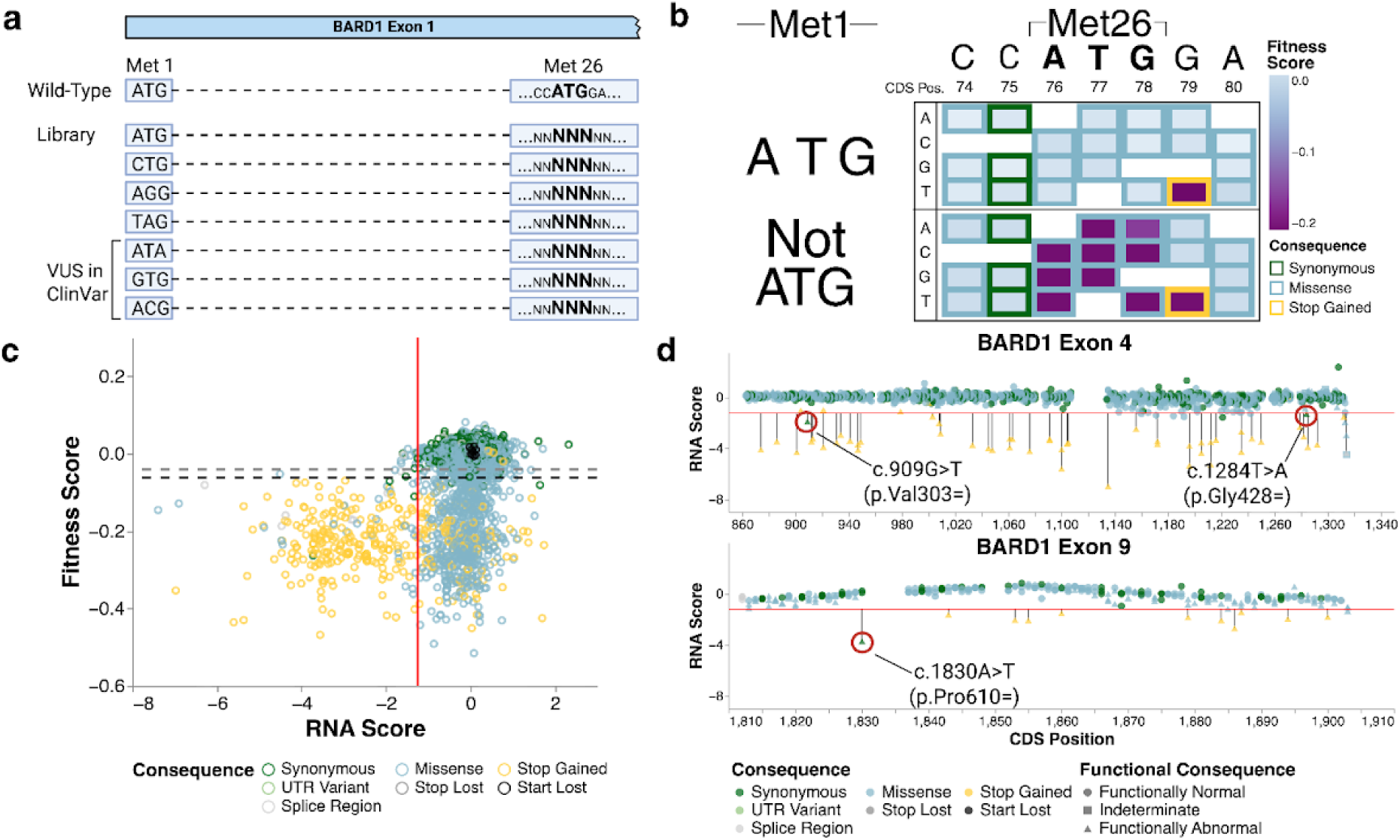
SGE identifies nucleotide variants that affect BARD1 expression and RNA abundance. (a) Design of double mutant experiments to investigate start codon usage. A repair template library for exon 1A was designed such that each variant a p.Met26 was coupled to either the reference ATG at p.Met1 or seven other codons including variants currently listed in ClinVar. (b) Fitness scores for double mutants experiments are displayed in nucleotide-level heatmaps grouped by whether p.Met1 was encoded by ATG or a variant codon. Each position in the coding sequence for p.Met26 and its surrounding bases are labeled on the X-axis. The nucleotide change is denoted on the Y-axis. Each box represents a SNV and the outline is colored by the mutational consequence, the fill color represents the SGE score (above) or average SGE score (below) for double mutants with variants at p.Met1. (c) Scatter plot of RNA scores (X-axis) vs. fitness scores (Y-axis) (n = 6,384). Horizontal black and gray dotted lines denote LoF and functionally normal SGE thresholds respectively. The vertical red line at X = −1.24 represents the RNA score threshold used to call variants depleted in the RNA. (d) Lollipop plots of exons 4 and 9 highlight synonymous changes that show LoF fitness scores, depleted RNA scores, and are predicted to disrupt splicing. The red horizontal line represents the RNA score threshold for calling variants depleted in the RNA.

We utilized mRNA abundance measurements to investigate variant effects on *BARD1* splicing and expression. Since alternative splice isoforms of *BARD1* have been reported^54^ and can impact RNA abundance measurements, we first investigated *BARD1* splicing in HAP1. RT-PCR on wild type *BARD1* RNA detected two previously identified alternate splice isoforms: one lacking exons 2 and 3 and one lacking exon 3^54^ (**Extended Data Fig. 9a-b**). Given their low abundance and frameshifts, we did not expect these alternate transcripts to significantly impact RNA abundance measurements for variants in exons 2 and 3. Thus, we designed primers to reverse transcribe, amplify, and sequence each edited exon from the canonical transcript (MANE transcript ENST00000260947.9) to identify and count variants. RNA scores were generated by taking the log_2_ ratio of the variant’s RNA abundance over its genomic DNA abundance (**Fig. 1a**, **Fig. 4c, and Extended Data Fig. 8**).

26 functionally abnormal missense variants had low mRNA abundance, suggesting they compromise BARD1 function at the mRNA level by altering splicing. 62% (16 of 26) were predicted by SpliceAI^54^ (SpliceAI delta score >= 0.2) reinforcing this conclusion. Additionally, three synonymous variants, c.909G>T (p.Val303=), c.1284T>A (p.Gly428=), and c.1830A>T (p.Pro610=), were classified as LoF based on SGE fitness scores and also had low mRNA abundance (**Fig. 4d**). Despite being located away from exon-intron junctions, SpliceAI predicted high splice disruption likelihood with delta scores > 0.70 for all three variants (**Supplementary File 1**). Notably, p.Gly428= has been identified in the human population at low frequency^48^. Similarly, we identified eight intronic SNVs with LoF fitness scores (**Supplementary File 1**). Of these, five variants were predicted by SpliceAI to disrupt splicing.

These results provide new insights into BARD1 expression, demonstrating that BARD1 translation can initiate without the annotated start codon and identifying SNVs that disrupt proper splicing. These findings highlight that nucleotide-resolution SGE with direct genome editing can accurately detect variant effects on both RNA expression and protein function.

### SGE resolves missense loss of function mechanisms

Having validated that the variant effects measured by SGE are relevant to BARD1’s function as a tumor suppressor (**Fig. 3c**), we sought to rationalize the impact of missense variants in BARD1 on the basis of known structural features. Here we investigated sequence-function relationships across BARD1’s RING, ARD, BRCT domains and compared them to the BRCA1’s RING and BRCT.

The structurally similar BRCA1 and BARD1 RING domains were comparably sensitive to missense changes (21.3%^10,31^ vs. 20.1% LoF, p = 0.4). The BARD1 RING showed high missense sensitivity at functionally critical positions: 55 of 57 missense variants at the eight C3HC4 Zn^2+^-coordinating residues were LoF (**Fig. 5a and Extended Data Fig. 10**), and residues forming the interior of the 4-helix bundle BRCA1-BARD1 interface were also intolerant to substitution^11^ (**Fig. 5b**), consistent with disruption of this critical interaction.

**Figure 5.**
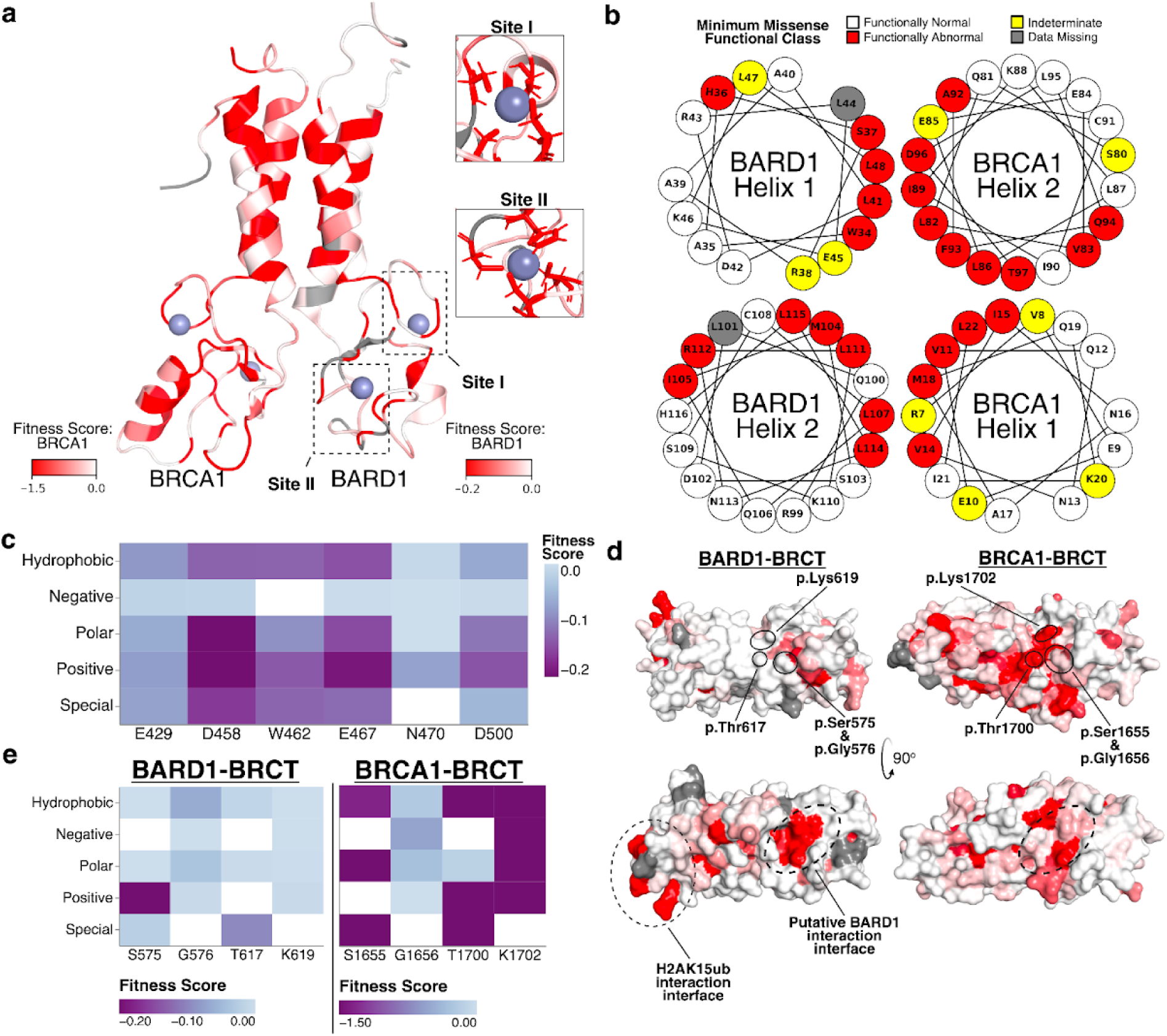
Structural consequences of BARD1 missense variants. (a) NMR structure of the BRCA1-BARD1 RING:RING dimer (PDB 1JM7^11^) colored by minimum missense SGE score for BARD1 or BRCA1^10,31^. Residues without fitness scores are colored gray. Residues in BARD1’s two zinc coordinating sites are highlighted. (b) The functional classes for variants in the 4-helix bundle of the RING domain are depicted as a helical wheel diagram. (c) Heatmap of median fitness scores for missense variants at BARD1 ARD residues reported to interact with the H4 N-terminal tail by type of amino acid introduced. Median fitness scores are colored as indicated. (d) Space-filling model of the crystal structures of the BARD1 BRCT (PDB 3FA2^68^, left) and the BRCA1 BRCT (PDB 1T29^69^, right) colored by median SGE score. Top orientation highlights the missense sensitivity of a lysine residue in the phosphopeptide binding site of both proteins. The bottom orientation highlights the H2AK15ub interaction interface of BARD1 and a missense sensitive surface on BARD1’s BRCT, hypothesized to be an interaction interface. (e) Heatmaps of median fitness scores for missense variants at residues in BARD1’s phosphopeptide binding site (left) and BRCA1’s phosphopeptide binding site (right) by the type of amino acid introduced. Fitness scores are colored for each protein as indicated.

The BARD1 ARD recognizes H4K20me0^12,14,25^, a marker of newly replicated chromatin that determines the cellular choice between NHEJ and HDR, while the BRCT enables rapid recruitment of the BRCA1-BARD1 dimer to dsDNA breaks^3,55^. Mapping minimum missense fitness scores onto the BARD1 ARD-BRCT-nucleosome structure revealed mutational intolerance at residues required for ARD’s interaction with the H4 tail. Residues forming an acidic pocket near H4K20me0 (p.Glu429, p.Asp458, p.Glu467) and a pocket for histone H4 p.His18 (p.Trp462, p.Glu467, p.Asn470, p.Asp500) were highly missense sensitive and did not tolerate positively charged substitutions (**Fig. 5c**). Similarly, BRCT-nucleosome residues p.Arg705 and p.Asp712 that form charged interactions with histone H2A and H2B respectively did not tolerate missense variants (**Extended Data Fig. 11**). Additionally, the BRCT recognizes DNA damage-dependent H2AK13ub/K15ub marks^19^. p.Gln715, which contacts Ub p.Thr66, showed the highest missense sensitivity (**Fig. 5d and Extended Data Fig. 11**). These results suggest that cell fitness and tumor suppression depend on BARD1’s ability to interact with nucleosomes and recognize H4K20me0 in newly replicated chromatin and DNA-damage-induced H2AK13ub/K15ub marks.

Furthermore, BRCT residues p.Ser575, p.Gly576, p.Thr617, and p.Lys619^55^ form a phosphopeptide binding motif. Missense substitutions introducing hydrophobic or negatively charged residues were tolerated—contradicting the hypothesis that BARD1 requires phosphopeptide binding for HDR^14,55^ (**Fig. 5d-e**). In contrast, the analogous motif in BRCA1’s BRCT (p.Ser1655, p.Gly1656, p.Thr1700, p.Lys1702), which binds a BACH1 phosphopeptide^56,57^, were intolerant to charge-disrupting variants measured by SGE^10,31^. This differential sensitivity indicates that unlike BRCA1, the function of BARD1 that supports cellular fitness is phosphopeptide-independent.

Moreover, this motif binds poly (ADP-ribose) (PAR) for BARD1’s role in stalled fork protection (SFP) and recruitment to sites of DNA damage^14,15,55^. However, tolerance of charge-disrupting variants like p.Lys619Glu demonstrates that PAR binding is dispensable for essential BARD1 function, suggesting BARD1’s HDR function alone is sufficient for HAP1 cell survival. With SGE accurately predicting cancer risk (**Fig. 3b**) and enrichment of LoF BARD1’s BRCT missense variants in breast cancer cases (**Fig. 3c**), we provide evidence that BRCA1 and BARD1’s role in HDR is required for tumor suppression in humans. This agrees with studies in mice where the murine equivalent to p.Lys619 was dispensable for HDR and tumor suppression^15,25,58^.

Comparing mutational impacts on BARD1 and BRCA1’s BRCT domains revealed comparable missense sensitivity (23.2% vs. 22.3% LoF^10,31^, p = 0.93), consistent with their similar structure. Despite this similarity, protein-specific variant impacts emerged. Beyond differential sensitivity at phosphopeptide binding sites, we identified a missense-sensitive acidic surface on BARD1’s BRCT (residues p.Glu648, p.Glu649, p.Glu665) absent in BRCA1’s BRCT (**Fig. 5d**). We hypothesized this surface is a binding interface for an unidentified, BARD1-specific interactor required for HDR. However, examination of predicted BARD1 interactions^59^ and attempts to dock BARD1 interacting proteins^22^ using AlphaFold-Multimer^60^ did not yield any credible interactions. Future work is required to investigate the role of this pocket in BARD1 function.

Altogether, our identification of functionally critical BARD1 residues demonstrates that BARD1’s role in HDR is critical for tumor suppression, highlighting SGE’s ability to link specific cellular functions to disease.

## Discussion

Here we applied SGE to measure the impact of 95% of all possible SNVs and 3-bp deletions in *BARD1* on cell survival and RNA expression. SGE discriminated damaging variants, such as nonsense and canonical splice variants, from likely benign variants like synonymous and intronic changes. Most importantly, SGE enabled functional classification of missense and splice region variants with uncertain effects, providing critical evidence for clinical variant classification.

To validate that our SGE assay captures disease-relevant biology, we demonstrated that LoF missense variants overall, as well as those within the ARD and BRCT domains, are significantly enriched in breast cancer cases compared to controls. The choice of HAP1 cells is further supported by recent work comparing *BRCA1* SGE data in HAP1 cells (myeloid origin) versus human mammary epithelial cells^31^ (HMEC). The results indicate that HAP1-derived fitness scores more accurately predict variant effects in human cancer than HMEC-derived data, possibly because haploid HAP1 cells are uniquely sensitive to loss of BRCA1-BARD1-mediated HDR activity.

The high sensitivity and specificity of our SGE results enabled calibration to strong evidence levels for both pathogenic and benign classifications using the OddsPath framework^50^. Achieving strong evidence calibration required including nonsense variants in the pathogenic truth set, as ClinVar contains only one confirmed pathogenic missense variant in BARD1. Our demonstration that LoF missense and nonsense variants are enriched similarly in breast cancer cohorts shows both confer similar disease risk, supporting this approach, which has precedent in *BAP1*^34^, *BRCA1*^61^, and *RAD51C*^36^.

LoF missense variants were predominantly confined to the three functional domains (RING, ARD, BRCT), consistent with their established roles. However, the comprehensive nature of SGE revealed important exceptions that challenge structure-function models. Our programmed 3-bp deletion scan showed that almost no single amino acid in the exon 4-encoded disordered domain is required for BARD1 function, suggesting considerable flexibility. In contrast, the missense variant, p.Lys140Asn, previously shown to be critical for catalyzing end resection^62^ and RAD51 loading to promote HDR^42^, scored as LoF (c.420C>G) or indeterminate (c.420C>A), in our assay despite the tolerance of most individual 3-bp deletions in this region. This suggests that multiple positively charged residues and local flexibility, rather than specific residues, are key functional requirements.

SGE also revealed that start-loss variants and nonsense variants upstream of p.Met26 retained function, indicating the annotated start codon is dispensable. Our double-mutant experiments provide definitive evidence that either p.Met1 or p.Met26 can independently support full BARD1 function and normal RNA expression. These findings have direct clinical implications: it is highly likely that start-loss variants at p.Met1 that are currently classified as VUS and the nonsense variant p.Glu19Ter, currently classified as likely pathogenic in ClinVar, are benign. We have reclassified start-loss VUS c.3G>A and p.Glu19Ter, which were in Ambry’s dataset, to LB given our functional evidence (**Supplementary Table 2**).

We have generated strong functional evidence for the pathogenicity and benignity for 4,541 missense variants and show that LoF *BARD1* variants increase cancer risk. This enables reclassification of 95.4% (1,382 of 1,488) *BARD1* variants for which we could apply functional evidence, representing 87.6% of VUS catalogued by Ambry Genetics. Beyond hereditary breast and ovarian cancer, LoF *BARD1* variants have been associated with neuroblastoma^63^, where PARP inhibition is effective therapy^18^. Our study immediately increases the utility of clinical genetic testing targeting *BARD1*—both for informing patients of cancer risk and guiding treatment.

## Methods

### sgRNA design and cloning

All sgRNAs were designed with Benchling’s “CRISPR” function using default settings. Off- and on-target scores for all guides were generated using GRCh38. For most targets, two sgRNAs were selected based on ability to make a synonymous change to the protospacer/PAM, minimal predicted off-target effect, and maximum predicted on-target effect. For synonymous changes to the PAM site, C>G and G>C changes were preferred. If no guides had PAM sites amenable to a synonymous change, two synonymous changes under the protospacer were made to prevent recutting.

All sgRNAs were ordered from Integrated DNA Technologies (IDT) and cloned according to the protocol deposited here: http://dx.doi.org/10.17504/protocols.io.3byl46ddjgo5/v1.

sgRNA sequences were confirmed by Sanger. Sequence-verified plasmids were propagated overnight in 150 mL LB and purified with ZymoPure II Maxiprep kits andEndoZero spin columns (Zymo Research) (**Supplementary File 2**).

### Repair template library design and cloning

The SGE target sequence (∼120 bp) was obtained from GRCh38 and the MANE Select transcript `ENST00000260947.9`. In addition to fixed edits at the sgRNA binding site, a neutral substitution was introduced at an alternative CRISPR cut site to allow for use of an alternate sgRNA if necessary. Otherwise, a second synonymous substitution was made on the opposite side of the SGE target sequence to mark successful HDR. The SGE target sequence annotated with fixed edits and HDR markers served as template to program all possible SNVs and 3-bp deletions. 3-bp deletions were designed to start at every nucleotide, some span two adjacent codons. The double-mutant library used the target sequence and fixed edits of library 1A as the original template. At the canonical start codon, six alternate codons plus the original methionine were designed and used to program double-mutant repair templates comprising all possible SNVs spanning bases c.74 to c.80. For all targets, an additional ∼20 bp of genomic context upstream and downstream of the sequence were appended to act as PCR handles.

Programmed sequences were ordered as pooled oligonucleotides and array-synthesized (Twist), except for the double-mutant library, which was ordered as an oligonucleotide pool from IDT. Oligonucleotides corresponding to a specific target were amplified from the oligo pool by qPCR (Kapa HiFi, Kapa Biosystems) using target-specific PCR handles. Reactions were terminated before plateau and products were cleaned using Sera-Mag Select beads (Cytiva).

Homology arms for each target were PCR-amplified (Kapa HiFi) from wild-type HAP1 gDNA using primers targeting sequences 1,000 to 1,500 bp up- and downstream of the target. A subsequent PCR added adaptors for Gibson Assembly into pUC19 digested with EcoRI (NEB) and SalI (NEB). The homology arms and pUC19 plasmid were assembled using NEBuilder HiFi Assembly (NEB), transformed into Stellar Competent Cells (Takara), and plated on media containing ampicillin. Colonies were miniprepped (NEB), and sequence-confirmed (Plasmidsaurus). To assemble the homology arms with the repair template library amplified from the pooled oligonucleotides, sequence-confirmed homology arm pUC19 plasmids were linearized using region specific primers by inverse PCR (Kapa HiFi), digested with DpnI (NEB), and gel extracted (NEB) prior to assembly using NEBuilder HiFi Assembly (NEB). Assembly reactions were cleaned using Clean and Concentrator (Zymo Research) and transformed into electrocompetent 10-beta cells (NEB). After transformation, 1% was plated on ampicillin plates to assess efficiency. A minimum of 10,000 colonies were targeted, representing approximately 20X coverage of the library. The remaining 99% was used to start a 150 mL overnight culture, from which libraries were purified using ZymoPure Maxiprep Kits and EndoZero spin columns and sequence-confirmed.

### Cell culture and time point sampling

Cas9-HAP1 Δlig4 cells were obtained from Horizon Discovery. Cells were cultured in Iscove’s Modified Dulbecco’s Medium (IMDM) (Gibco) supplemented with 10% fetal bovine serum (MilliporeSigma). For all SGE experiments, cells were routinely sorted for 1N haploid population^10^, expanded, and frozen until use.

For each SGE target, cells were thawed into four 15 cm plates (Genesee Scientific) five days prior to transfection at a density of 5 million cells per plate. Cells were resuspended into 25 mL of media containing 10 μg/mL of blasticidin (Gibco) to select for Cas9 expression. Four days prior to transfection, the blasticidin-containing media was refreshed. Two days later, cells were taken off blasticidin and seeded the following day into 10 cm plates (Genesee Scientific) at a density of 8 million cells per plate, with each plate used for one transfection.

Cells were transfected using Xfect (Takara) according to the manufacturer’s protocol. For each transfection, 12 μg of sgRNA and 3 μg of HDR library were used. For negative control transfections, 3 μg of HDR library was used with 12 μg of sgRNA targeting housekeeping gene *HPRT1*. For each SGE region, ten transfection reactions were prepared: 1 negative control and 9 replicates. Transfected plates were incubated at 37°C for at least 4 hours, then the transfection reagent-containing media was aspirated and fresh media added after 2x 10 mL washes with phosphate buffered saline (PBS) (Gibco). Plates were incubated overnight at 37°C.

One and two days after transfection, the media was swapped for media containing 10 μg/mL blasticidin and 3 μg/mL puromycin to select for cells actively expressing Cas9 and successfully transfected with the sgRNA plasmid. Blasticidin and puromycin media was replaced with antibiotic-free media three days after transfection.

Days 5, 9, and 13 after transfection, cells were sampled and passaged. Samples taken on days 5 and 13 were used for sequencing. On day 5, three transfection plates were combined to yield three replicates. 50% was collected for DNA and RNA extraction and 50% was replated in antibiotic-free media and incubated at 37°C until day 9. On days 9 and 13, 80% cells were sampled and 20% passaged.

### Sequencing preparation and sequencing

gDNA and RNA were isolated from day 5 cell pellets using AllPrep DNA/RNA kits (Qiagen). From day 13 pellets, only gDNA was extracted with DNeasy kits (Qiagen). gDNA and RNA yields were quantified using Qubit 1x Broad Range Kit (Invitrogen) and NanodropOne (Invitrogen) respectively.

Sequencing libraries were prepared with three sequential PCR steps (Kapa HiFi). First, eight 50 μL reactions were prepared for each day 5 replicate and 16 for each day 13 replicate; all reactions contained 250 ng of template. PCR was performed using the minimum number of cycles as determined from a previous qPCR reaction using wild-type HAP1 gDNA as template.

To allow for selective amplification from HAP1 gDNA and not residual plasmid background, primer pairs were designed such that one primer annealed outside of the homology arm and the other within the homology arm. After amplification, 10 μL of the respective reactions for each sample were pooled and cleaned using Sera-Mag Select beads (Cytiva).

Next, Illumina-specific adapters were added to cleaned PCR products with qPCR.. Repair template library preparation also began at this step, using 2 ng of repair template. Products were cleaned by Sera-Mag Select beads, A final qPCR added sample-specific sequencing indices. All reactions were monitored and removed from the thermal cycler prior to plateau.

For each sample, up to 5 μg of isolated RNA from day 5 pellets was reverse-transcribed (Superscript IV, Invitrogen) using a primer specific to *BARD1*’s 3’ UTR, and treated with RNase H (NEB). cDNA sequencing library preparation involved either two or three qPCRs. For exons >300 bp in length, three qPCRs (Kapa HiFi) were performed due to sequencing read length limitations. The first qPCR selectively amplified the targeted exon from synthesized cDNA using primers in flanking exons. Eight 25 μL replicate reactions were performed for each sample. 5 μL from each replicate were pooled and cleaned using Sera-Mag Select beads. In the subsequent reaction, only the region of interest was amplified using target-specific primers (**Supplementary File 2**) that included adaptors for Illumina sequencing. Products were cleaned by Sera-Mag Select beads and used for a final indexing qPCR. For exons <300 bp in length, two qPCRs were performed. The first qPCR was analogous to the first qPCR for large exons; however, primers were designed to include Illumina-specific adaptors. After pooling, products were cleaned by Sera-Mag Select beads and indexed in a subsequent qPCR.

Cleaned, indexed PCR products were quantified using Qubit 1x High-Sensitivity Kit (Invitrogen). Libraries were pooled and sequenced on the Illumina NextSeq2000 platform.

### Processing of sequencing data

Paired-end reads were adapter-trimmed and merged with `SeqPrep` (version 1.3.2) with the following options: `-A GGTTTGGAGCGAGATTGATAAAGT -B CTGAGCTCTCTCACAGCCATTTAG -M 0.1 -m 0.001 -q 20 -o 20.` Reads containing N bases were discarded. Merged reads were mapped to target-specific, GRCh38 reference sequences using the `mem` algorithm in `bwa` (version 0.7.17-r1188) to produce target-specific BAM files.

Counts of each programmed SNV within each replicate and at each timepoint (henceforth, “dataset”), and within the repair template library, were extracted from each BAM file by a custom Python script. All mismatches between the merged read sequence and the reference sequence were identified. Reads were considered valid if they had the expected length, contained all fixed edits, and had a single additional mismatch within the edited region of the target, which was taken as the programmed edit. Additional mismatches within the merged read but falling outside of the edited region were tolerated. Filtering was relaxed for SGE targets containing homopolymer runs with length ≥4. In such cases, a change in sequenced homopolymer length of up to two was allowed.

Genomic coordinates harboring a cell line variant in HAP1 cells were discarded from analysis. Programmed variants with fewer than ten observations in the repair template library or any dataset were excluded from analysis.

Counts of each programmed 3-bp deletion within each dataset, and within the repair template library, were similarly extracted from BAM files by a custom Python script. CIGAR strings were parsed to identify alignments containing a single 3-bp deletion in the merged read sequence. Reads containing such deletions were further examined to ensure that all fixed edits were present, except for those that overlapped with the identified, programmed deletion. Reads containing sequence-level mismatches within the edited region as well as a single programmed deletion were discarded; sequence mismatches outside of the edited region were tolerated.

Each variant’s count within each dataset was converted to a variant frequency by dividing the integer count by the number of valid reads within the dataset.

The editing rate per dataset was calculated by dividing the sum of valid SNV reads and valid deletion reads by the total number of reads.

### Quality control

Several quality control metrics were computed for all sequenced libraries associated with each SGE target. For the starting library, at least 500,000 merged reads were required, of which at least 30% were required to be valid and no more than 10% could be wild-type. The negative control library could contain no more than 1% valid reads. Each replicate dataset was required to have at least 150,000 valid reads. At least two valid replicates were required for each timepoint. Finally, the Pearson correlation coefficient was calculated between each pair of replicates within each timepoint, and a minimum value of no less than 0.5 was required. Library 4J failed quality control and was removed from the dataset.

### Smoothing

A frequency ratio was computed for each variant in each day 5 replicate, comprising the log_2_ of its frequency at day 5 over its frequency in the repair template library. A one-dimensional LOESS smoother, implemented in Python with the `loess`^64^ package (version 2.1.2) with the option `span=0.20`, was used to attenuate the presumed position effect on the observed variant counts owing to the location of the cut-site relative to the programmed edit. The smoother was fit separately within each replicate to the log_2_ ratios at day 5 to yield fitted log_2_ ratios. These positional fits were then subtracted from the day 5 log_2_ frequency ratios, and from each replicate’s cognate day 13 log_2_ frequency ratios, to yield adjusted log_2_ ratios. The model was fit only on those variants whose frequencies at day 5 were no less than half of their starting frequencies.

### Scoring

A continuous-time linear regression model, implemented in the `linregress` module of `scipy`^65^ (version 1.14.0), was used to estimate SNV and deletion scores as a function of the adjusted log_2_ frequency ratios within each replicate and each timepoint: 0, 5, and 13 days. The log_2_ variant frequency at the starting timepoint was set to 0. Estimated log_2_ fold-changes per unit time (i.e., per day) were taken as the fitness score for each variant. For variants covered by two adjacent SGE targets, frequencies from both targets were jointly modeled to produce a single score. For the double mutant library, fitness scores were estimated through applying a continuous negative binomial model, implemented through `PyDEseq2`^66^, to variant counts at each timepoint.

RNA scores were generated for variants in the cDNA by calculating the log_2_ frequency ratio of the variant in the RNA to its respective day 5 DNA frequency. Since the genomic region sequenced in DNA samples is larger than the region sequenced in RNA samples, prior to RNA scoring, day 5 DNA frequencies were adjusted to include only positions sequenced in RNA samples. RNA scores were independently calculated for each experimental replicate and collapsed into a final score by taking the median of all replicate scores. For variants covered by two adjacent SGE targets, the final reported score is the mean of both individual scores.

### Variant annotation

The expected molecular consequence(s), HGVS identifiers, and amino acid change of each programmed variant were annotated by the Ensembl Variant Effect Predictor (`vep`)^67^, version 115, using the MANE Select *BARD1* transcript (`ENST00000260947.9`). For variants with multiple predicted molecular consequences, only the consequence with the highest predicted impact was retained.

### Functional class assignment

A two-component Gaussian mixture model, implemented in `scikit-learn` (version 1.7.1), was fit to the SNV scores. The model’s initial component means were set to the mean of the middle 95% of the score distribution of expected-neutral SNVs—specifically, synonymous and intronic SNVs—and the mean score of all nonsense SNVs outside of exons 1, 4, and 11. The probability of belonging to each component was estimated for each SNV, and those whose probability of assignment to a component exceeded 0.95 were assigned that component’s label: “functionally abnormal” and “functionally normal” for the components with the lesser and greater means, respectively. Remaining SNVs were labeled “indeterminate.” The approximate score threshold values for functional class assignment were identified by taking the scores of the variants that minimized the distance to one of the 0.95 probability cutoffs. These two score thresholds were then applied to the 3-bp deletion scores, without refitting, to yield the same three functional classes. The final fitness score thresholds for the functionally abnormal and functionally normal classes were −0.0615 and −0.0406.

Functional classes for RNA scores were determined using the mean and standard deviation of the bottom 97.5% of nonsense variant RNA scores, excluding exons 1, 2, and 11. The score threshold for “normal” RNA abundance was −1.244 and determined by calculating one standard deviation above the mean nonsense RNA score.

### Variant filtering

Programmed SNVs in the same codon as a suspected cell line SNP or a fixed edit required for SGE library design were scored; however, variant annotations based on the reference *BARD1* transcript may assign incorrect molecular consequences to these altered codon contexts. Consequently, these variants were removed from the final data set (**Supplementary File 1**). 15 variants: c.1851G>A, c.1851G>C, c.1851G>T, c.1850G>A, c.1850G49G>C, c.1850G>T, c.1849T>A, c.1849T>C, c.1849T>G, c.1727C>A, c.1727C>G, c.1727C>T, c.1726C>A, c.1726C>G, and c.1726C>T were required for structure-function analyses in Figure 5. Annotations for these variants were manually reviewed and corrected prior to analysis.

### ClinVar analysis

All SNVs and 3-bp deletions were accessed from ClinVar^9^ on September 12, 2025 and had at least 1-star review status.

### Case control analysis

Variant counts from the CARRIERS^8^ and BRIDGES^6^ breast cancer case-control cohorts were received from Dr. Fergus Couch or downloaded from https://tinyurl.com/BRIDGESSummary, respectively. Variants were lifted over to GRCh38 coordinates. The total number of patients and controls were retrieved for each cohort from their respective publications. Additionally, both cohorts reported the number of individuals from studies that did not select patients on the basis of family history (population-based) and studies that were enriched with patients with a family history of breast cancer. Odds ratios were calculated for both BRIDGES and CARRIERS cohorts individually considering all individuals and only considering individuals from population-based studies. For the CARRIERS population-based cohort only, an additional analysis was conducted subsetting for estrogen receptor (ER) negative breast cancer cases only. Training variants for the REVEL and MutPred-2 variant effect predictors were removed from the odds ratio calculation. Lastly, joint analyses were conducted by combining both BRIDGES and CARRIERS cohorts. All odds ratios were calculated using Fisher’s Exact tests.

### Variant reclassification

In order to obtain the appropriate strength of evidence for the functional data, two distinct functional evidence calibrations using the current ClinGen-approved calibration method were used to generate Bayesian likelihood ratios supporting PS3/BS3 criteria^46^. First, values were derived using missense, stop-gained, splice site and synonymous variants as control sets (**Supplementary Table 2**) which corresponded to strong strength of evidence for both benign (0.0054) and pathogenic variants (316.23). Additionally, OddsPath values were recalculated using only missense variants as controls. Functional data revealed strong strength of evidence for pathogenic variants (18.5), whereas the OddsPath for benign variants was indeterminate (0.514) (**Supplementary Table 2**).

Clinically observed missense, nonsense, synonymous and intronic variants in *BARD1* were obtained from Ambry Genetics, along with publically available data on population frequency (gnomAD v4.0), *in silico* predictors (BayesDel and SpliceAI), previous functional studies^39,40^, and PVS1 annotations with thresholds applied by Ambry’s internal classification scheme for *BARD1* for variant interpretation. The dataset comprised 1,720 total variants: 1,578 variants of uncertain significance (VUS), 101 likely pathogenic or pathogenic (LP/P) variants, and 41 likely benign or benign (LB/B) variants.

For reclassification, functional data were applied to variants in conjunction with existing computational, population frequency, segregation, and de novo evidence, and 161 variants had indeterminate or no functional data (**Supplementary Table 2**). ACMG/AMP v3 points based thresholds were used for the classifications. A manual review was performed by a genetic counselor and a molecular pathologist to verify the accuracy and consistency of the initial and final variant classifications.

### External data sources

SpliceAI^54^ variant annotations were annotated by the Ensembl Variant Effect Predictor (`vep`). ClinVar^9^ data for germline SNVs and deletions with at least a 1-star review status were downloaded on September 12, 2025. Variants annotated as “Benign/Likely benign” were grouped with “Likely benign” variants and variants annotated as “Pathogenic/Likely pathogenic” were grouped with “Likely pathogenic” variants. Variants classified with “Conflicting interpretations of pathogenicity” were grouped with variants annotated as “Uncertain significance”. Allele frequencies for SNVs were downloaded from gnomAD^48^ v.4.1.0 on September 5th, 2024 (https://gnomad.broadinstitute.org/) and from the Regeneron Million Exome Variant browser^49^ v1.1.3 (https://www.rgc-research.regeneron.com/me/home) on August 2nd, 2024. Claude (Anthropic) was used to refine text, and figures were made using Biorender.

## Resource Availability

### Lead contact

Further information and requests for resources and reagents should be directed to and will be fulfilled by the lead contact, Lea M. Starita

### Materials availability

Please contact the corresponding author with requests for any reagents generated by this study.

### Data and code availability

SGE bioinformatics pipeline:

https://github.com/bbi-lab/sge-pipeline/

Supplementary files, supplementary tables and code to regenerate all figures and analyses: https://github.com/ivanw314/BARD1_SGE_analysis

Raw sequencing reads can be downloaded from IGVF: https://data.igvf.org/analysis-sets/IGVFDS4537CHCJ/

Fitness scores are available through MaveDB: https://www.mavedb.org/score-sets/urn:mavedb:00001250-a-1

### Ethics statement

Use of de-identified human sequence data was approved by UW IRB study #3598.

## Data Availability

All data, supporting files, and software are available online.

https://github.com/bbi-lab/sge-pipeline/

https://github.com/ivanw314/BARD1_SGE_analysis

https://data.igvf.org/analysis-sets/IGVFDS4537CHCJ/

https://www.mavedb.org/score-sets/urn:mavedb:00001250-a-1

## Acknowledgements

We thank current and former members of the Starita lab, the Brotman Baty Advanced Technology Lab, the IGVF Coding Variant Focus Group, the ClinGen/AVE Functional Data Working Group, and Rachel Klevit for helpful discussions. We thank Fergus Couch for sharing variant frequencies from the CARRIERS study. We are grateful to the Brotman and Baty families for their generous gift establishing the Brotman Baty Institute for Precision Medicine, and to the scientific and administrative directors Jay Shendure and Nola Klemfuss for their support in making this work possible. This work was funded by the Brotman Baty Institute, NIH NHGRI IGVF Consortium (5UM1HG011969), GREGoR Consortium (5U01HG011744), Advancing Genomic Medicine Research Consortium (R01HG013025), and Centers of Excellence in Genome Sciences (5RM1HG010461). R.K.G. acknowledges support from the Washington Research Foundation postdoctoral fellowship. A.E.M. was supported by an Early Career Award from the Alex’s Lemonade Stand for Childhood Cancer and RUNX1 foundation (21-25037), and the Brotman Baty Institute Catalytic Collaborations Grant (CC28).

## Author Contributions

This project was conceptualized by I.W., S.C., and L.M.S. The original manuscript was drafted by I.W., P.G., and L.M.S. The manuscript was reviewed by all authors and revised by I.W., S.C., M.W.S., N.T.S., M.T., P.G., A.E.M., A.H., A.X., R.K.G., T.B., M.E.R., S.P., D.M.F., and L.M.S. Experiments were designed by I.W., S.C., M.D., S.F., and L.M.S. Experiments were performed by I.W., S.C., N.T.S., S.B., M.P., A. Ha., A.H., and A.X under the supervision of L.M.S. Experimental data was analyzed and visualized by I.W., M.W.S., and P.G., with feedback from D.M.F, and L.M.S. Clinical variant interpretation was led by P.G., M.T., and A.E.M., with support and feedback from T.B., M.E.R., and L.M.S. Overall project administration was handled by S.H. and L.M. Funding was acquired by D.M.F. and L.M.S.

## Extended Data Figures

**Extended Data Figure 1.**
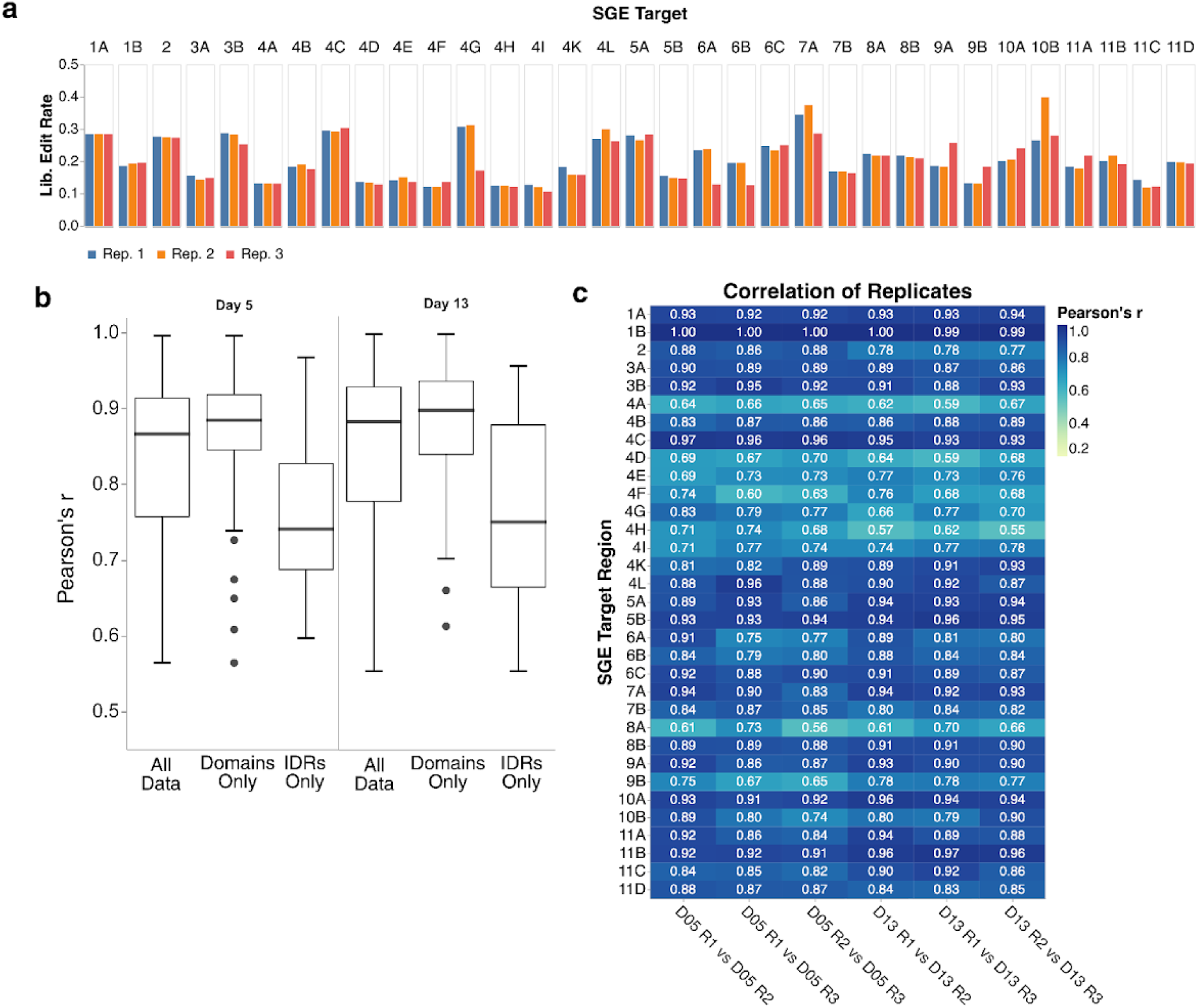
Editing rates and reproducibility across SGE experiments. (a) Bar charts showing editing rates (Y-axis) generating usable reads across all QC-passing targets (Y-axis) and replicates (bar color). Usable reads are defined as having all fixed edits and a single programmed SNV or 3-bp deletion. (b) Box and whisker plots of median Pearson’s r correlation (Y-axis) across day 5 (left) and day 13 (right) timepoints. Data was subsetted as described on the X-axis. “All data” represents all data, “Domains Only” represents data only from the 3 structured domains, “IDRs Only” contains data from the exon 4-encoded disordered domain. (c) Heat map of Pearson’s r correlation between replicates (X-axis) for all QC-passing targets (Y-axis). Pearson’s correlation is colored for each target and replicate comparison.

**Extended Data Figure 2.**
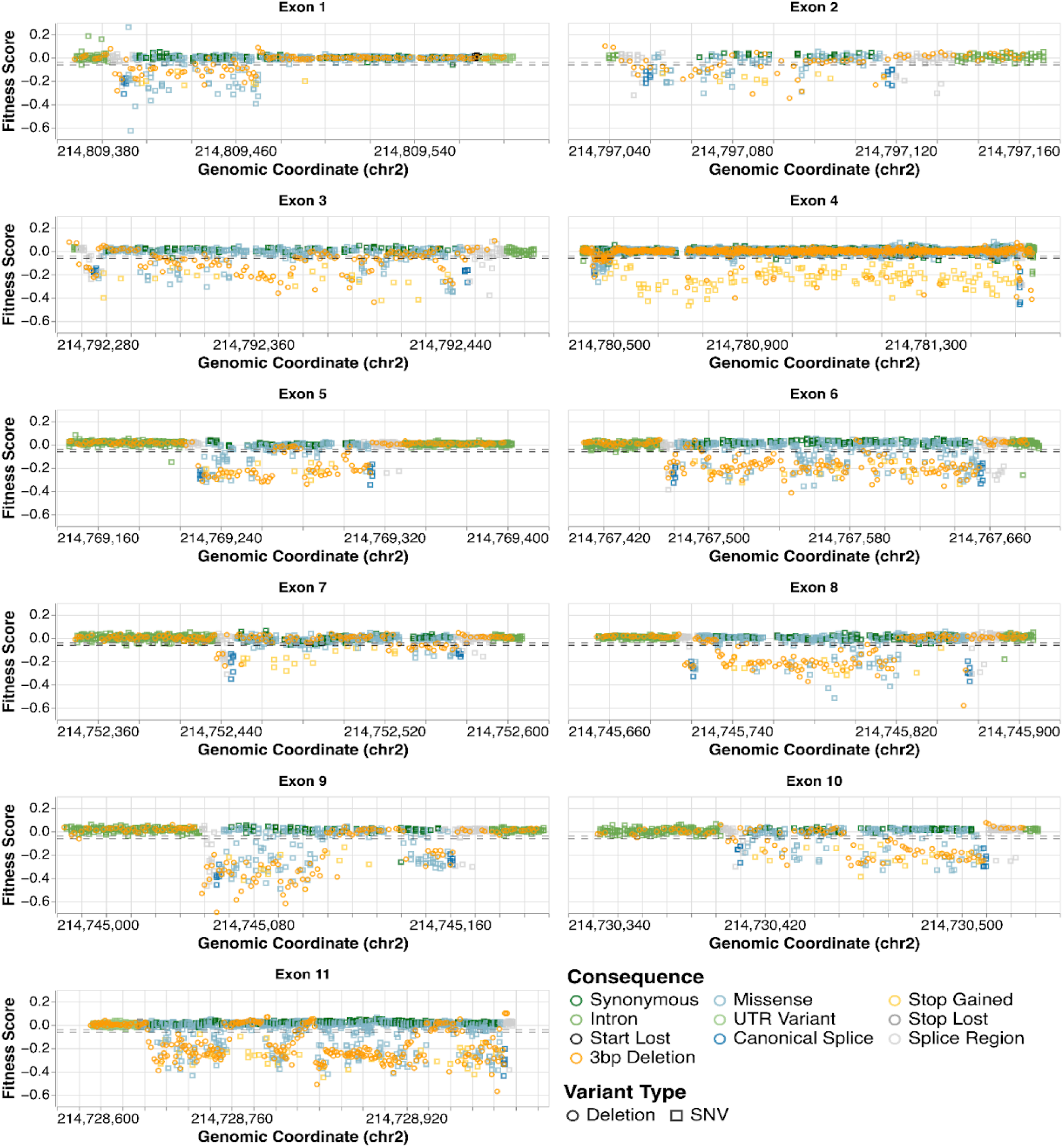
Fitness scores for all SNVs and 3-bp deletions across *BARD1* exons. Scatterplots of SGE score (Y-axis) vs. 1-based, hg38 genomic coordinates (X-axis) for each *BARD1* exon. SNV and inframe deletion variant types are represented by shape and molecular consequence of the variant is represented by color. Horizontal dotted red and blue lines represent score thresholds for classifying variants as LoF or functionally normal respectively.

**Extended Data Figure 3.**
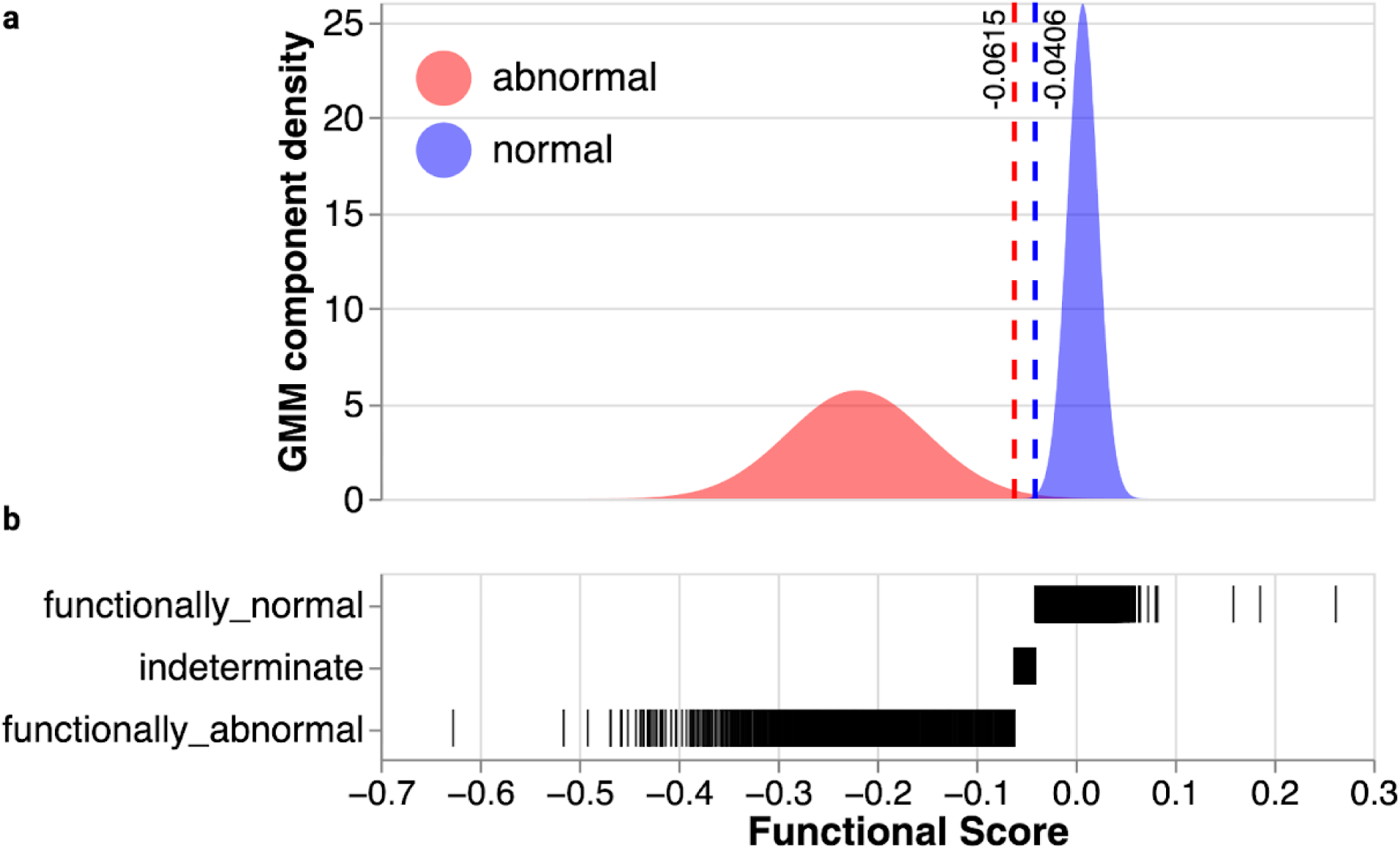
Fitted gaussians for variant functional classification. (a) “Abnormal” (red) and “Normal” (blue) gaussians used to draw thresholds for classifying variants as LoF or functionally normal. Estimated density from GMM-modeling is on the Y-axis and SGE score is on the X-axis. Vertical red dashed line at X = −0.0615 represents upper threshold for LoF variants. Vertical blue dashed line at X = −0.0406 represents lower threshold for functionally normal variants. (b) Strip plot of functional class (Y-axis) vs. SGE score (X-axis)

**Extended Data Figure 4.**
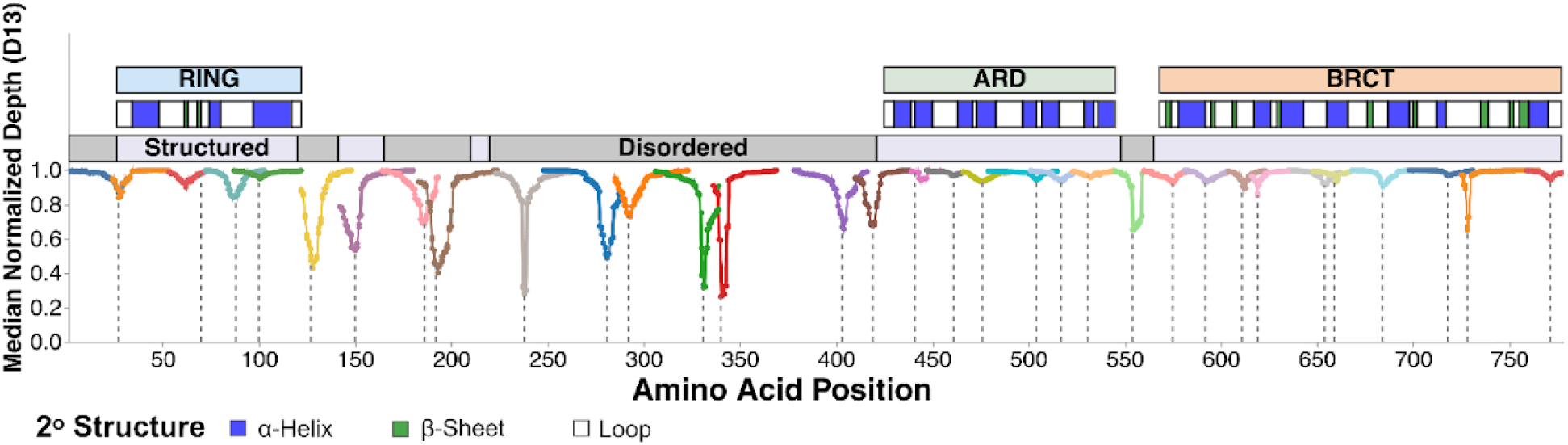
Median day 13 normalized read depth across *BARD1*. Line plot of minimum median day 13 normalized depth (Y-axis) for all bases contributing to a single amino acid residue (X-axis) across the protein coding sequence of *BARD1*. Minima represent residues where sequencing reads contain deleted bases. Line color denotes different SGE targets. Vertical dashed gray lines denote CRISPR-Cas9 cut sites. Above the plot, the top-most track represents the BARD1 domains. The middle track represents secondary structure elements of each domain (white - loop, blue - alpha-helix, green - beta-sheet) as seen in solved structures for the RING (PDB 1JM7^11^), ARD (PDB 3C5R^13^), and BRCT (PDB 3FA2^68^). Bottom-most track depicts if the region is predicted to be structured (pLDDT > 50) (lavender) or unstructured (gray) by AlphaFold2^70^.

**Extended Data Figure 5.**
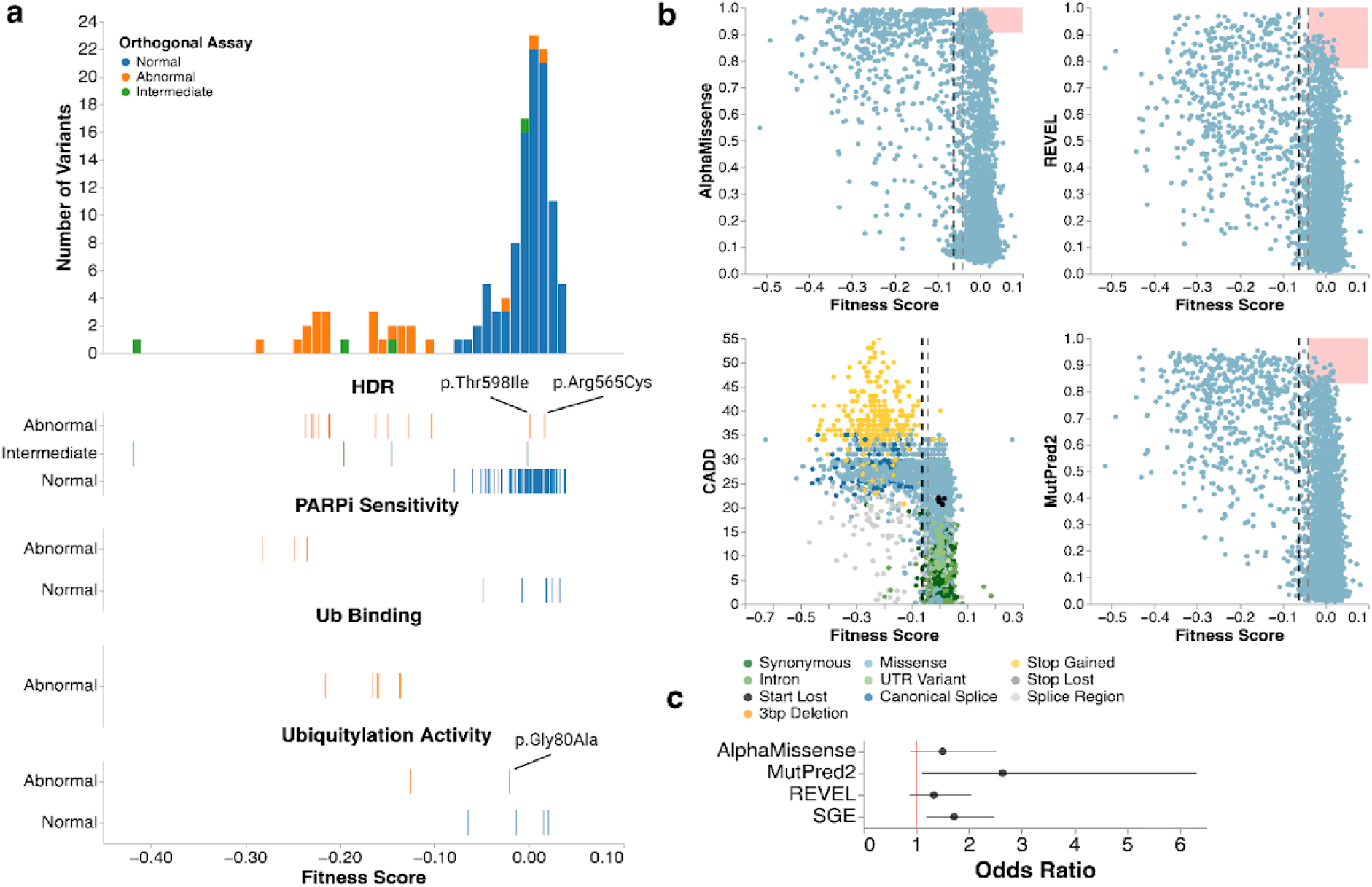
Comparison of BARD1 fitness scores to variant effect predictors and orthogonal BARD1 functional assays. (a) Histogram (top) and strip plots (bottom) of 125 variants assayed in orthogonal functional assays vs. SGE score (X-axis). In all plots, color represents the functional consequence of the variant in its respective assay. In the strip plots, plots are titled by the orthogonal assay used and the Y-axis represents the functional consequence of the variant. 3 variants that were functionally abnormal in orthogonal assays but functionally normal in SGE are highlighted. (b) Scatter plots comparing AlphaMissense^43^ (top-left), REVEL^45^ (top-right), CADD^47^ (bottom-left), and MutPred-2^44^ (bottom-right) to fitness scores. Predictor scores are on the Y-axis and fitness scores on the X-axis. Variants are colored by molecular consequence. Vertical red and blue lines represent SGE score thresholds for classifying variants as LoF or functionally normal respectively. Overlaid red rectangles highlight regions where predictors overpredict pathogenicity. Rectangles are drawn using the estimated score thresholds to yield moderate+ pathogenic evidence for each predictor^46^. (c) Odds ratios (X-axis) for occurrence of missense variants predicted to be LoF by predictors or SGE (Y-axis) in breast cancer cases vs. healthy controls. Results generated from the combined BRIDGES^6^ and CARRIERS^8^ population-based cohort. The vertical red line denotes an odds ratio of one. Points denote the estimated odds ratio and whiskers denote the 95% confidence interval.

**Extended Data Figure 6.**
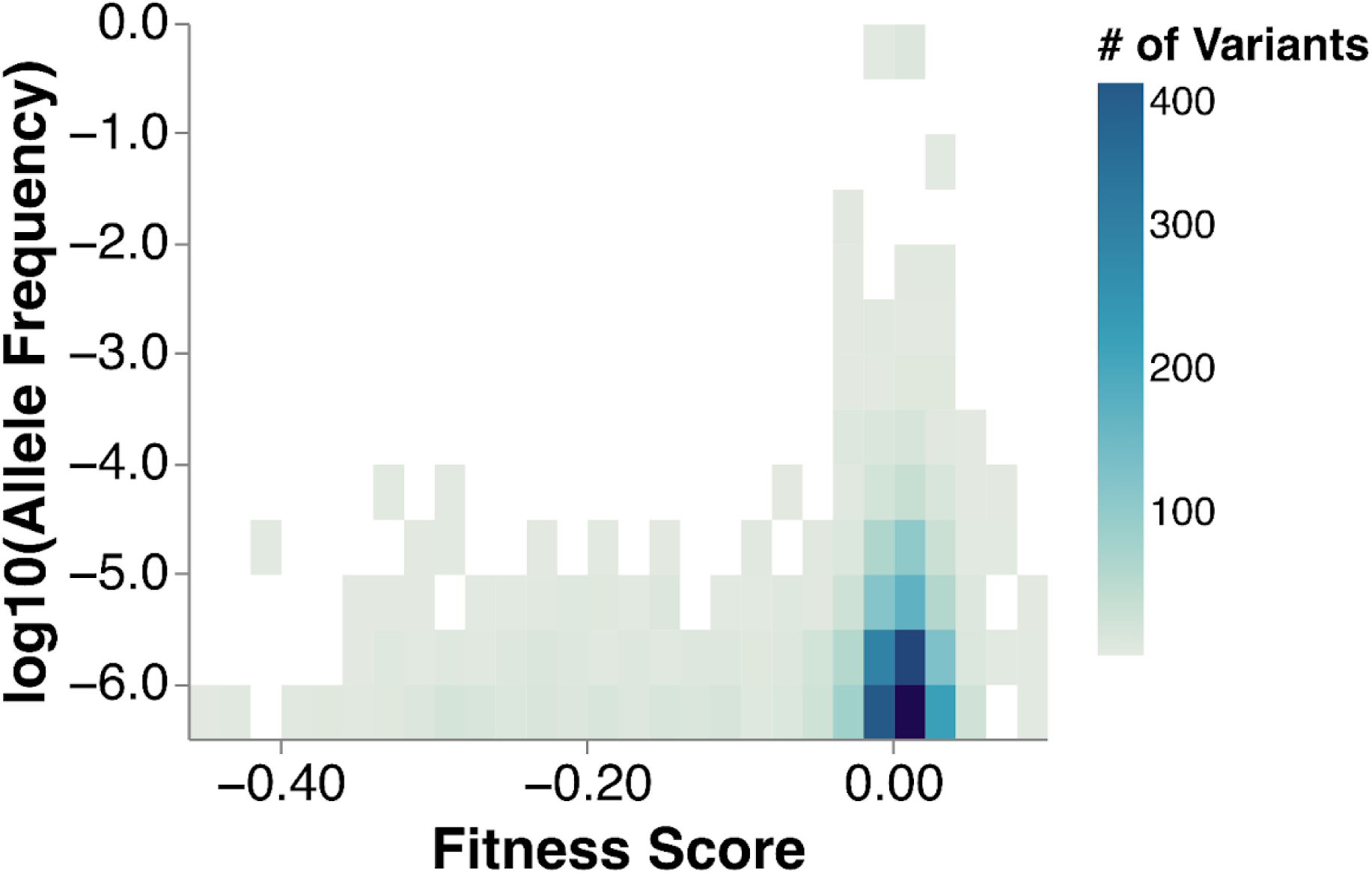
Mean allele frequency of *BARD1* variants assayed by SGE. Heatmap of fitness scores (X-axis) compared to log_10_-transformed minor allele frequency (MAF) (Y-axis) (n = 3,435). Variants were accessed from gnomAD^48^ and the Regeneron Million Exome Variant Browser^49^. Each bin is colored by the indicated number of variants in that bin.

**Extended Data Figure 7.**
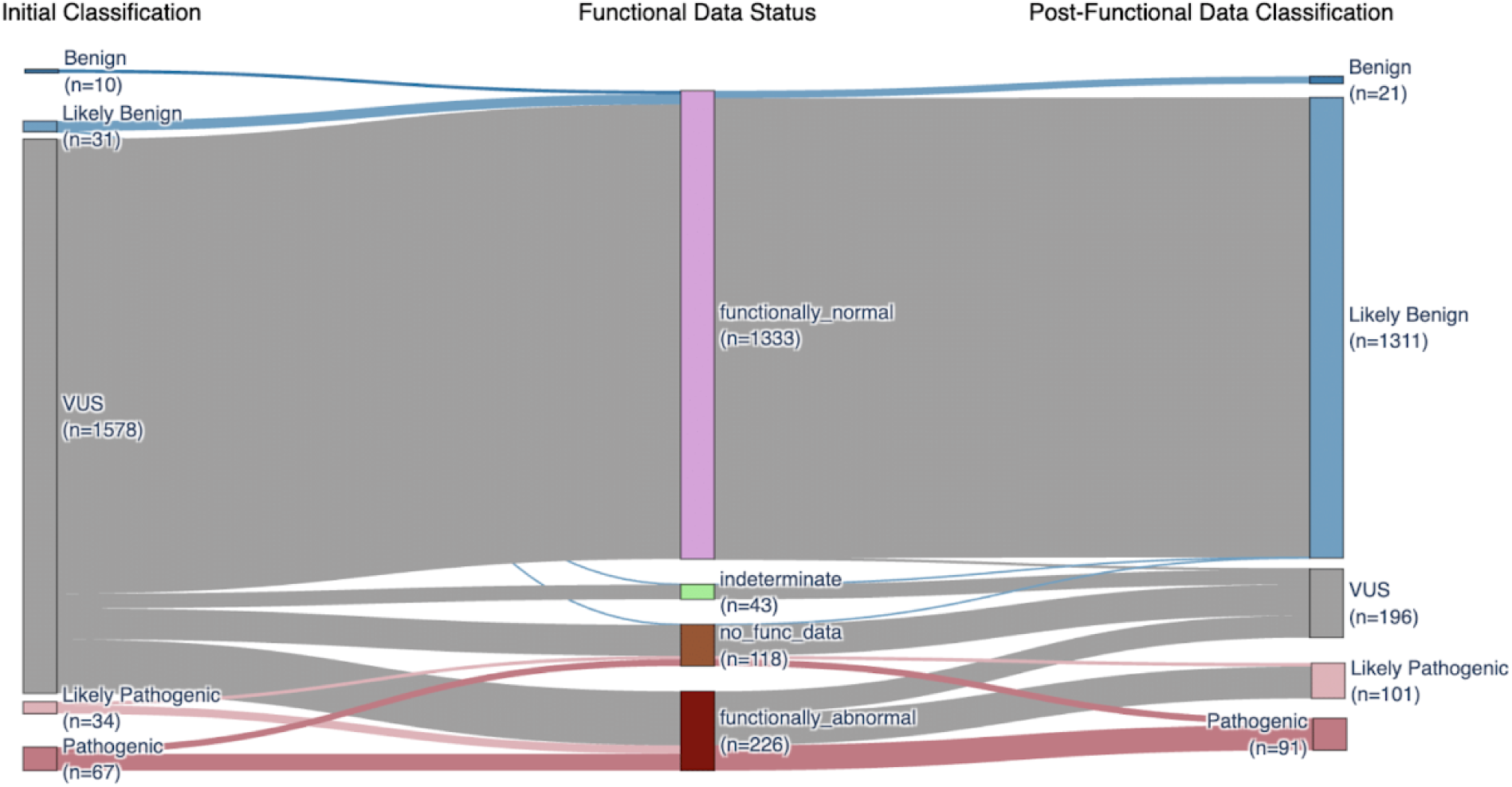
The clinical utility of data generated by *BARD1* SGE. Sankey plot of all variants in Ambry Genetics’ database before and after application of functional data as indicated.

**Extended Data Figure 8.**
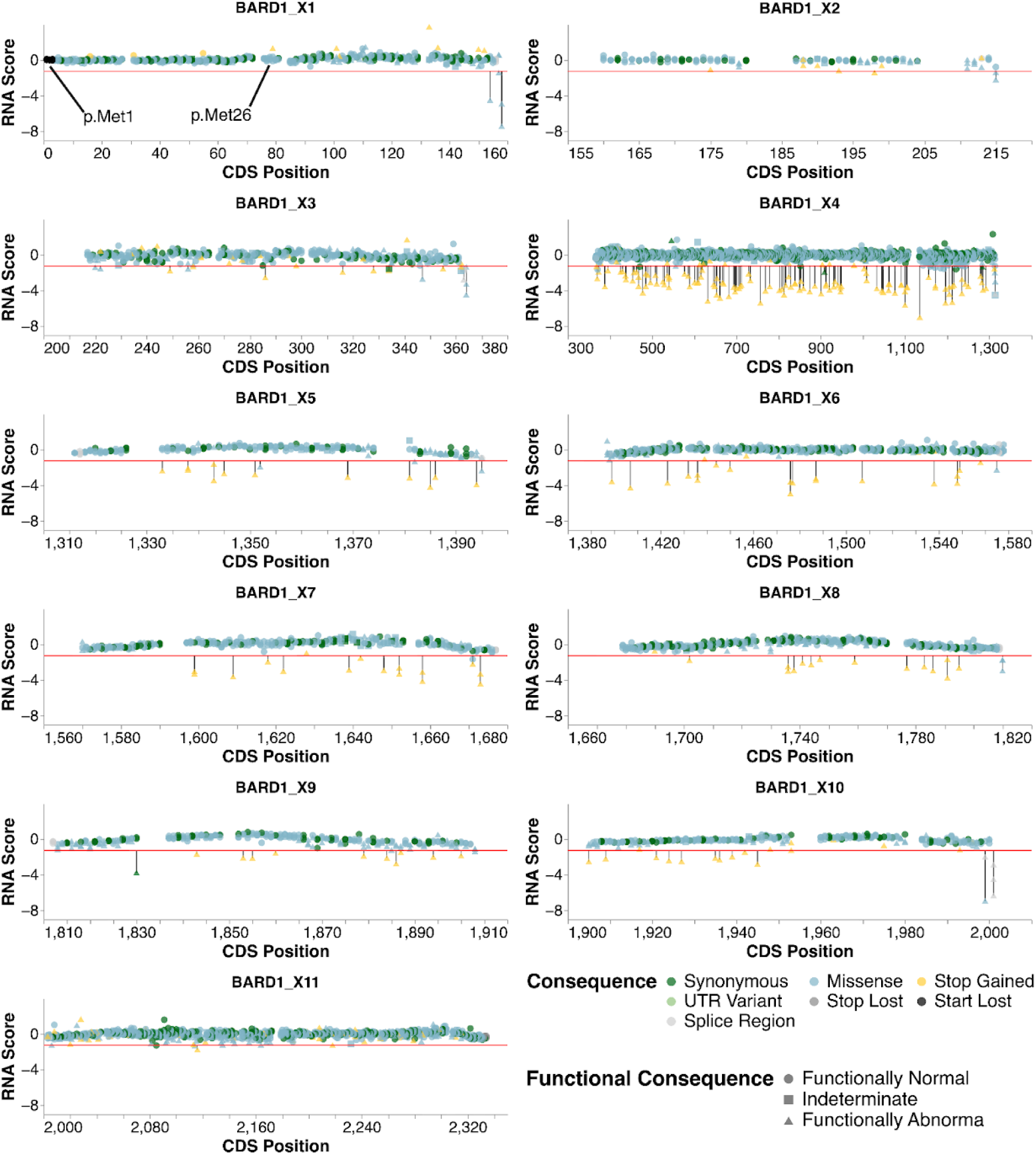
RNA scores for variants across *BARD1*. Scatter plots of RNA score (Y-axis) vs. CDS position (X-axis) for all exons in *BARD1*. For all plots, the horizontal red line at Y = −1.24 represents the RNA score threshold used to classify variants as having “low” RNA abundance, representing one standard deviation above the mean RNA score for nonsense variants. Variants are colored by molecular consequence and shape denotes the SGE functional consequence. Variants classified as LoF with low RNA abundance are further highlighted with a black vertical line extending from the RNA threshold to the datapoint. In exon 1, variants stop-loss variants and variants impacting alternate start codon p.Met26 are highlighted.

**Extended Data Figure 9.**
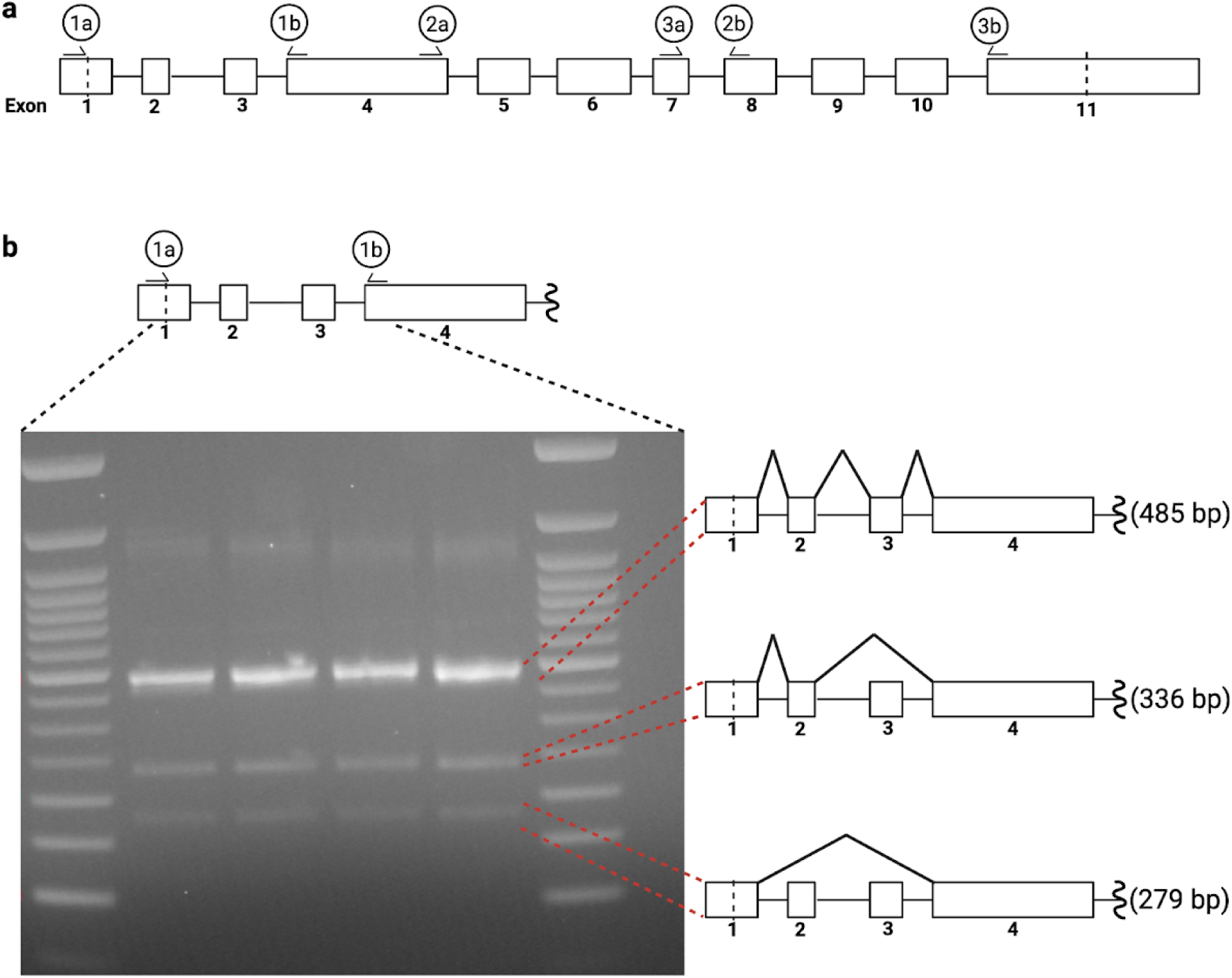
Alternate splicing of the *BARD1* transcript in HAP1. (a) Illustration of *BARD1* exons and primers used for RT-PCR to assess presence of alternate *BARD1* isoforms in HAP1. Large rectangles represent *BARD1* exons and are numbered below. Vertical dashed lines in exon 1 and 11 represent the start and stop codons respectively. Labeled half arrows above rectangles represent forward and reverse primers used for RT-PCR. Primer pairs are labeled by integer. Forward/reverse primers are denoted by a/b respectively. (b) Illustration in (a) zoomed to highlight the region amplified by primer pair 1 (top). Gel image of resultant RT-PCR on bottom left. 3 possible products are identified and the splicing events required to yield the respective product are depicted (right).

**Extended Data Figure 10.**
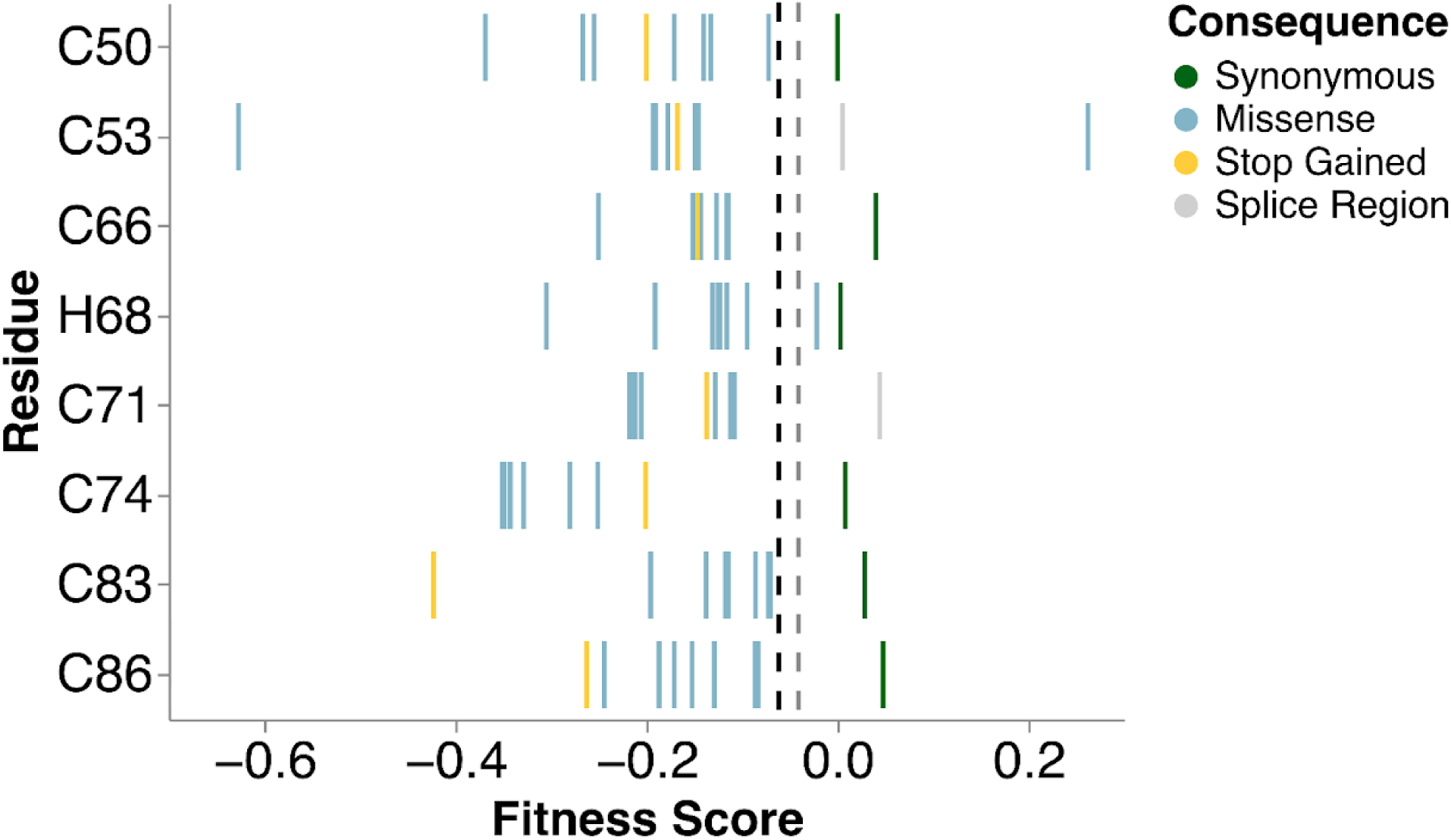
Fitness scores for missense variants at Zn^2+^ binding residues. Strip plot of fitness scores (X-axis) for all variants at the C3HC4 residues (Y-axis) colored by indicated mutational consequence (n = 57). Each Cys and His residue position is labeled. Vertical dashed black and grey lines indicate functional class thresholds.

**Extended Data Figure 11.**
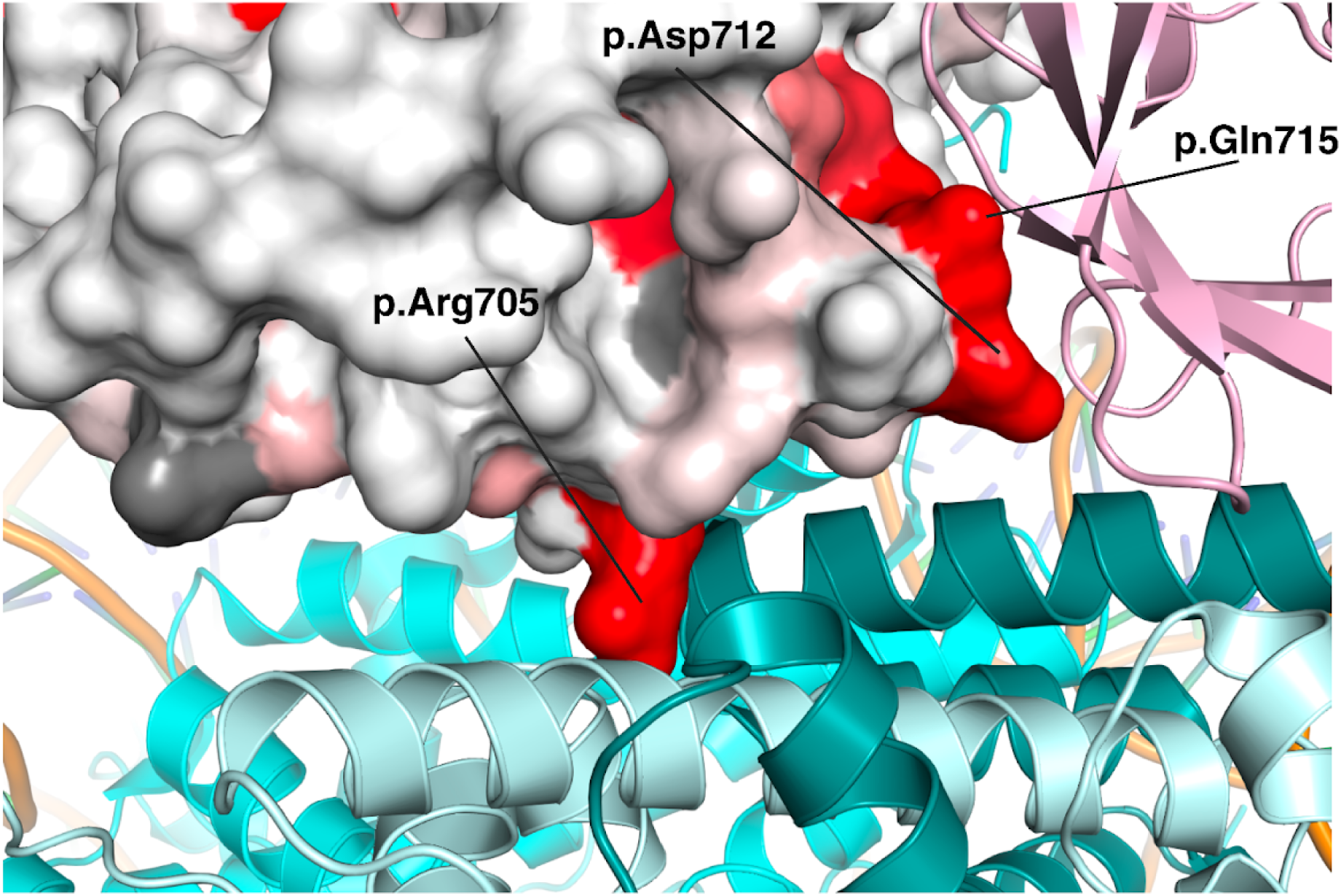
Consequences of BARD1 missense variants on BARD1 BRCT interactions. Crystal structure of BARD1 ARD and BRCT domains bound to the nucleosome core particle (NCP) and histone H4 (PDB 7LYC^14^) zoomed in to focus on the interactions between the BRCT, histones H2A/H2B (light blue/dark teal) and ubiquitin (pink). A space-filling model represents the BRCT with the surface colored by mean missense score. Residues reported to be interacting are labeled.

